# The role of physical activity programmes in mitigating obesity and type 2 diabetes in Singapore using simulation-based forecasts until 2050

**DOI:** 10.1101/2025.08.14.25333249

**Authors:** Bekzod Normatov, Haolong Song, Shihui Jin, Xinyu Zhang, Yichen He, Nigel WH Lim, Vong Fee Zheng, Muhammad Hafiz Bin Mohd Aziz, Falk Müller-Riemenschneider, Charmaine Pei Ling Lee, Borame L Dickens

## Abstract

**Background:** The National Steps Challenge (NSC) is a physical activity intervention that has enrolled over 2.1 million people in Singapore across 9 years. The impacts of this intervention on current and future obesity, and type 2 diabetes mellitus (T2DM) prevalence are currently unknown. Here, we estimate population-level impacts and explore future scenarios in how physical activity interventions could mitigate rising obesity and T2DM burdens.

**Methods:** Using a microsimulation model of 7.9 million Singapore residents and NSC participation data, we modelled the impacts of physical activity on BMI trajectories and T2DM prevalence across the Singaporean population from 1990 to 2050 by ethnicity, sex, age, and body-mass-index subgroups. We then simulated five programme scenarios, ranging from no NSC to full population-wide deployment, projecting estimated future impacts.

**Findings:** Without the NSC, total obesity prevalence was projected to rise from 13.6% (total n=261,675) in 1990 to 32.4% (total n=1,189,081) by 2050, and T2DM from 5.8% (total n=111,597) to 16.0% (total n=587,200). Based on current recruitment and programme retention rates, relative to the No NSC, total obesity and T2DM prevalence could decrease by 15.8% and 6.2%, respectively, by 2050. These reductions are estimated to vary substantially by ethnicity and sex, and programme scenario.

**Interpretation:** Physical activity programmes such as the NSC can help mitigate rising obesity and T2DM prevalence when implemented at the population level, as seen in Singapore. However, their success depends on robust digital infrastructure and sustained incentivisation to enhance long-term impact.

## Introduction

High body mass index (BMI) contributes to approximately 5 million deaths annually from non-communicable diseases.^1,2^ An estimated 1 in 8 people worldwide are living with obesity, conventionally defined as having body mass index (BMI) ≥ 30 kg/m^2^, and by 2030 this ratio is projected to reach one in five for women and one in seven for men.^3,4^ Moreover, 43% of adults are estimated to be overweight, and by 2035, this number is projected to reach 51% of the global population.^5,6^ Associated complications from obesity include type 2 diabetes mellitus (T2DM), where obese individuals are at seven times higher risk of T2DM compared to those at normal weight.^7^ Thus, the global prevalence of diabetes is also rising with a projected 10·1% increase from 2020 to 2030.^8^

Within Asia, the risk of developing obesity-related complications starts at a lower BMI, which necessitates a lower obesity cut-off at 27·5 kg/m^2^.^9,10^ Moreover, with more than 60% of the world’s diabetic population living in Asia, the global rise in diabetes disproportionately affects this region.^11^ This trend is particularly pronounced in South-East Asia, where projections suggest a concerning 16·3% rise in diabetes prevalence from 2014 to 2030.^8^ In response, government programmes have been designed that target modifiable lifestyle factors where physical activity (PA) programmes have shown substantial benefits among its participants.^12,13^ WHO has also developed the Global Action Plan on Physical Activity 2018-2030 that aims to achieve a 15% reduction in global physical inactivity by 2030 based on evidence of its impact on physical health.^14^

When the PA amount is of sufficient magnitude to produce an energy deficit of around 500-700 kcal per day, substantial weight loss can occur which has been observed in multiple randomised control trials (RCTs).^15^ In an RCT study of 54 obese women, participants who performed brisk walking or light jogging on a treadmill until they expended 500 kcal per day for 14 weeks decreased their body weight by 6·1 kg (∼6·5% of initial weight).^16^ Similar and consistent results have also been observed for obese men.^17^ In the Midwest Exercise Trial 2, an RCT study of 141 overweight and obese men and women in the US, an average significant weight loss of 3.9 kg and 5·2 kg was observed in the two treatment groups, where participants exercised at either 400 kcal/session or 600 kcal/session, five days/week, for ten months.^18^ In a similar study, STRRIDE, those who engaged in 27·2 km of walking each week (approximately 5000 steps per day) for eight months observed an average body weight loss of four kg (SD=3·5) without a decrease in caloric intake.^19^ Limited information exists however on the population-wide benefits of PA interventions, their long-term effects on health outcomes and how much they could mitigate the rising prevalence of obesity and diabetes.

In Singapore, in response to increasing obesity (22·3% in 2022) and type two diabetes (11·4% in 2024) prevalences, the National Steps Challenge (NSC) was introduced in 2015 as a large-scale population health intervention to promote physical activity.^20,21^ Since its inception, it has recruited over 2·1 million participants across eight waves. The NSC is a large-scale health initiative, aiming to increase physical activity levels among its participants through daily step and activity tracking. Participants are provided with free wearable devices and incentivised with rewards for meeting step-count goals.

As one of the largest physical activity interventions globally at a population level, we aim to estimate its long-term effects on BMI trajectories and T2DM prevalence outcomes, and project these potential benefits to 2050 under different recruitment scenarios using an individual-based simulation model of 7·9 million people with detailed sociodemographic, BMI trajectory, and diabetic status data. The effectiveness of the current NSC is evaluated by comparing projected outcomes without the NSC to alternative scenarios of enhanced recruitment and sustained retention. This modelling approach enables us to quantify the programme’s potential to reduce future chronic disease burden and inform policy decisions around scaling up national-level physical activity interventions.

## Methods

### Data

The analysis uses National Steps Challenge (NSC) data, which is anonymised, de-identified and includes 2·1 million unique NSC participants’ demographic, health, and physical activity measures across time. Launched by the Health Promotion Board (HPB), the NSC started in 2015, targeting Singapore residents aged 17 and above, with the introduction of NSC1, which attracted a total of 129,693 participants. NSC was then conducted annually and split into multiple waves (NSC1, NSC2, NSC 3, NSC4, NSC5, NC6) with an additional bonus round in 2021 (NSC5·5) and the final introduction of the NSC Always-on (NSCAO) in 2022. At its peak, during the waves of NSC3, NSC4, and NSC5, the number of active participants, who clocked at least 1000 daily steps, averaged at 527,000 per challenge year. Each NSC wave was split into a six-month challenge period and six-month off period where participants can enrol on a voluntary basis. During the challenge period, participants are incentivised to record their step counts and physical activity on their own tracker devices or a free NSC wearable tracker to gain Healthpoints, which could be redeemed for vouchers.

We calculated the primary outcome measure as the annual change in BMI (ΔBMI) using annually recorded height and weight records from the NSC participants. We excluded participants whose height and weight data were missing or extreme (i.e., height < 100 cm, height > 220 cm, weight < 25 kg, weight > 300 kg). Demographic characteristics, including sex, age, and ethnicity, were retrieved from participants’ national records upon registration.

### Microsimulation Model

Our microsimulation model tracks the resident population of Singapore from 1990 to 2050 (7·9 million people) for single-year age groups and three major ethnicities (Chinese, Indian, Malay) (Figure S1). Fertility rates by ethnicity were forecasted using Bayesian Structural Time Series models.^22^ We used annual fertility rates for each ethnic group during their respective ‘low fertility phases’.^23^ These phases began when fertility rates dropped below 2·1 for two consecutive years -starting from 1975 for Chinese and Indians, and from 2003 for Malays, all extending to 2022. We applied a logarithmic transformation to capture the slowing rate of fertility decline during these phases.^24^ 5,000 Markov chain Monte Carlo (MCMC) draws were generated for each ethnic group, with their means serving as point estimates (Figure S4). We used age-specific averaged mortality rate data from 2016 to 2019 to exclude any variations caused by COVID-19 for mortality projections as rates have remained stable during this period (Figure S5). As migration has also remained relatively stable, we assumed that the distribution of migrants across age would remain constant (Figure S6). We constructed a hierarchical model to simulate individual BMI trajectories across adulthood by sex and ethnicity, using longitudinal and national survey data, while T2DM risk was modeled using a logistic regression framework, where the annual probability of disease onset for each individual was specified as a function of their simulated BMI trajectory, age, sex, and ethnicity.^20,21^ Once an individual becomes a sufferer of T2DM, they are included within diabetes prevalence, even if the disease is well-managed. Further methodological details on these models are provided in the Supplementary Information 1 (Figures S2, S3, S7).

### Estimating physical activity levels

We used participants’ daily step count data to model physical activity patterns across demographic groups. Step count data were aggregated at the weekly level to account for daily variability. Each individual enrolled in the NSC had a baseline step count, as well as a change in step count during the intervention phase. Baseline step count was calculated by averaging each participant’s daily non-zero step counts during the 11-week pre-intervention period (prior to the incentive phase in NSC3, NSC4, and NSC5). We included only those participants with a baseline step count of at least 1,000 steps, resulting in a final sample of 120,575 participants. Baseline step count values varied by sex, ethnicity, age, and BMI group (72 strata). We modelled baseline step counts using gamma distributions with hyperparameters (shape and scale) shared among individuals within the same demographic stratum (Figure S8). Shape and scale parameters for each stratum were estimated using Maximum Likelihood Estimation (MLE) (Table S2).

We obtained each participant’s change in step count by subtracting their baseline step count from their daily non-zero step count, averaged across the intervention period (weeks 12–37 in NSC3, NSC4, and NSC5). We included only those actively enrolled participants who recorded a weekly average of at least 1,000 daily steps over the 26-week challenge period, resulting in a total sample of 66,734 participants. As with baseline step counts, we modelled the change in step count using gamma distributions across 72 demographic strata. Shape and scale parameters for each stratum were estimated using Maximum Likelihood Estimation (MLE) (Figure S9, Table S3).

### NSC Participation Model

Using NSC data, we developed three participation models to represent distinct annual participant statuses: first-time participants, continuing participants, and dropouts. These models operate alongside the longitudinal BMI model to simulate individual BMI trajectories over the lifespan. For first-time participants, we applied a Bayesian generalised linear model (GLM) with an identity link to predict first-year BMI change, incorporating baseline BMI, age, sex, ethnicity, baseline step count, and change in step count as covariates (n=60,238; see Supplementary Information 2). For continuing participants, those who re-enrolled in subsequent NSC seasons, we used a similar Bayesian GLM framework to estimate the effect of physical activity on annual BMI change (n=25,966; Supplementary Information 2). In both models, posterior parameter distributions were obtained using 5,000 MCMC samples, with posterior means used as point estimates.

Participants were classified as dropouts if they either did not enrol in a subsequent NSC season or averaged fewer than 1,000 steps per day over the 26-week intervention period. Among 2,944 dropouts, we observed a mean annual BMI increase of 0·142 units, consistent with estimates reported in the literature.^25–27^ If a participant re-enrolled after dropping out, they were treated as a first-year participant in their subsequent enrolment.

For each NSC season, we quantified the enrolment rate (the proportion of new participants relative to the eligible population), retention rate (the proportion remaining in the programme from one year to the next), and dropout rate (the proportion exiting the programme), as summarised in Table S4.

### Simulation Scenarios

To comprehensively evaluate the long-term public health impact of the NSC we simulated five distinct scenarios, each reflecting a different level of programme implementation and uptake, with six-month on and six-month off periods consistent with NSC (Table 1).

**Table 1.** Enrolment rate, retention rate and rationale for five simulation scenarios covering years 2022-2050.

| Simulation Scenario | Enrolment rate (%) | Retention rate (%) | Rationale |
| --- | --- | --- | --- |
| *No NSC* | 0 | 0 | Serves as a reference scenario to quantify the underlying burden of obesity and T2DM in the absence of any further population-wide physical activity intervention, enabling rigorous assessment of the incremental benefits of NSC. |
| *Average NSC* | 9·6 | 28·3 | Represents the expected impact of maintaining the NSC programme at mean historical enrolment and retention rates (NSC3–5), reflecting a realistic projection of current practice. |
| *Peak NSC* | 11·7 | 32·4 | Captures the programme’s maximum observed rates (NSC1–5) by modelling outcomes at the highest historical enrolment and retention rates, illustrating the effects of optimal past implementation. |
| *Double NSC* | 19·2 | 56·6 | Assesses the potential impact of substantially scaling up the programme through intensified policy and community efforts, helping to estimate the benefits of ambitious but potentially achievable expansion. |
| *Full NSC* | 100 | 100 | Provides an estimate of the theoretical maximum public health benefit by modelling universal participation, thereby defining the upper limit of preventable disease burden achievable through physical activity interventions. |

### Impact of the NSC on Obesity and Diabetes

Using the BMI and T2DM models, we estimated strata-specific prevalence of obesity and type two diabetes mellitus (T2DM) for each simulation scenario from 1990 to 2050. Obesity was defined using Asian-specific criteria (BMI ≥ 27·5 kg/m^2^). Each model iteration produced unique prevalence trajectories for both obesity and T2DM under the specified scenarios.

Results were aggregated across 1,000 simulation runs to generate median projected trends and corresponding simulation intervals.

### Sensitivity Analysis

We performed 1,000 runs of the DEMOS microsimulation model to generate a Monte Carlo sample, drawing parameters from posterior distributions to account for parametric uncertainty in the baseline population, BMI trajectories, and T2DM prevalence estimates. For each simulated population, obesity and T2DM prevalences were obtained by querying the model outputs. All analyses were conducted using R version 4·4·1.

### Ethics approval

Ethics exemption (review not required) was approved by the Department Ethics Review Committee of the Saw Swee Hock School of Public Health. This was based on the use of non-identifiable anonymised data.

## Results

### Obesity Prevalence

Under the baseline No NSC scenario, obesity prevalence among Singaporean adults aged 18 to 74 years is projected to rise from 13·6% (95% simulation interval: 12·8%–14·2%; total n=261,675) in 1990 to 32·4% (31·7%–33·4%; total n=1,189,081) by 2050. Among Chinese males, prevalence increases 11·1% (10·6%–11·9%; total n=84,537) in 1990 to 39·8% (38·8%–40·6%; total n=500,269). Malay and Indian male prevalences also demonstrate large increases, from 22·1% (21·4%–22·9%; total n=28,073) and 17·1% (16·2%–17·8%; total n=13,027) to 59·1% (58·1%–60·2%; total n=159,651) and 52·9% (51·4%–54·3%; total n=120,446), respectively. Chinese female prevalence shows a slight decline from 10·5% (9·9%–11·1%; total n=78,647) to 8·7% (8·2%–9·2%; total n=102,658), while Malay and Indian female prevalences show sharp increases from 31·3% (30·3%–32·2%; total n=38,991) and 25·8% (24·7%–26·7%; total n=16,225) to 52·6% (51·7%–53·7%; total n=129,206) and 66·3% (64·7%–67·5%; total n=123,081) respectively.

Under the Average NSC scenario, obesity prevalence among Chinese males is projected to reach 34·4% (28·8%–39·1%) in 2050, a 13·6% reduction (n=67,876) from baseline. Malay and Indian male prevalences are projected to reach 56·5% (52·3%–59·4%) and 48·3% (41·6%–53·5%), representing reductions of 4·4% (n=7,024) and 8·7% (n=10,474), respectively. Chinese female prevalence is projected to reach 6·7% (4·6%–8·5%) in 2050, a 22·9% reduction (n=23,600) from baseline. Malay and Indian female prevalences are projected to reach 48·0% (42·7%–52·6%) and 57·2% (50·2%–63·6%), reflecting reductions of 8·7% (n=11,299) and 13·7% (n=16,893), respectively.

The Peak NSC scenario results in only modest additional reductions compared to the Average NSC scenario, with declines ranging from 5·5% to 25·6% across groups. Under the Double NSC scenario, obesity prevalence among Chinese males is projected to reach 26·1% (15·6%– 35·8%) by 2050, a 34·4% reduction (n=172,203) from baseline. Malay and Indian male prevalences are projected to reach 53·5% (45·4%–58·9%) and 41·1% (26·7%–51·5%), corresponding to reductions of 9·5% (n=15,157) and 22·3% (n=26,867), respectively. Among females, prevalence generally stabilises. Chinese female prevalence is projected to reach 4·3% (1·5%–7·5%), a 50·6% reduction (n=51,919) from baseline. Malay and Indian female prevalences are projected to reach 41·3% (31·1%–49·9%) and 42·9% (30·0%–56·6%), reflecting reductions of 21·5% (n=27,757) and 35·3% (n=43,440), respectively.

The Full NSC scenario, which models universal participation among all adults aged 18 to 74 years, results in the most substantial declines. Obesity prevalence among Chinese males is projected to fall to 9·6% (2·4%–24·0%) in 2050, 1·5% lower than in 1990 and 75·9% lower (n=379,601) than baseline. Malay and Indian male prevalences are projected to reach 31·4% (14·6%–52·6%) and 20·0% (4·6%–43·5%), with reductions of 46·9% (n=74,828) and 62·2% (n=74,909), respectively. Chinese female prevalence is projected to reach 1·2% (0·0%– 3·7%), representing an 88·6% decrease from 1990 and an 86·2% reduction (n=88,498) from baseline. Malay and Indian female prevalences are projected to reach 14·8% (4·4%–32·6%) and 11·0% (4·1%–25·9%), reflecting reductions of 71·9% (n=92,851) and 83·4% (n=102,660) from baseline.

All NSC scenarios are projected to reduce obesity prevalence relative to baseline, but only the Full NSC scenario results in a clear reversal of trends across all sociodemographic groups.

### T2DM Prevalence

Under the baseline No NSC scenario, T2DM prevalence among Singaporean adults aged 18 to 74 years is projected to rise from 5·8% (95% simulation interval: 4·7%–7·6%; total n=111,597) in 1990 to 16·0% (13·3%–18·4%; total n=587,200) in 2050 (Figure 2). Among Chinese males, prevalence increases from 5·7% (3·9%–7·3%; total n=43,410) to 17·7% (12·7%–19·6%; total n=222,481) by 2050. Malay and Indian male prevalences have estimated increases from 7·5% (5·5–9·6%; total n=9,527) and 13·0% (9·9%–16·1%; total n=9,904) to 28·2% (24·1%–32·9%; total n=76,179) and 32·9% (28·3%–37·9%; total n=74,909), respectively. Chinese females show the lowest risk, with their prevalence increasing slightly from 4·7% (2·8%–6·1%; total n=35,204) to 7·4% (5·7%–9·2%; total n=87,318). Malay and Indian female prevalences undergo sharper increases, from 7·4% (5·3% to 9·6%; total n=9,218) and 8·7% (6·2%–10·9%; total n=5,471) to 20·6% (17·0%– 23·9%; total n=50,602) and 31·5% (26·9%–36·7%; total n=58,477), respectively.

**Figure 1.**
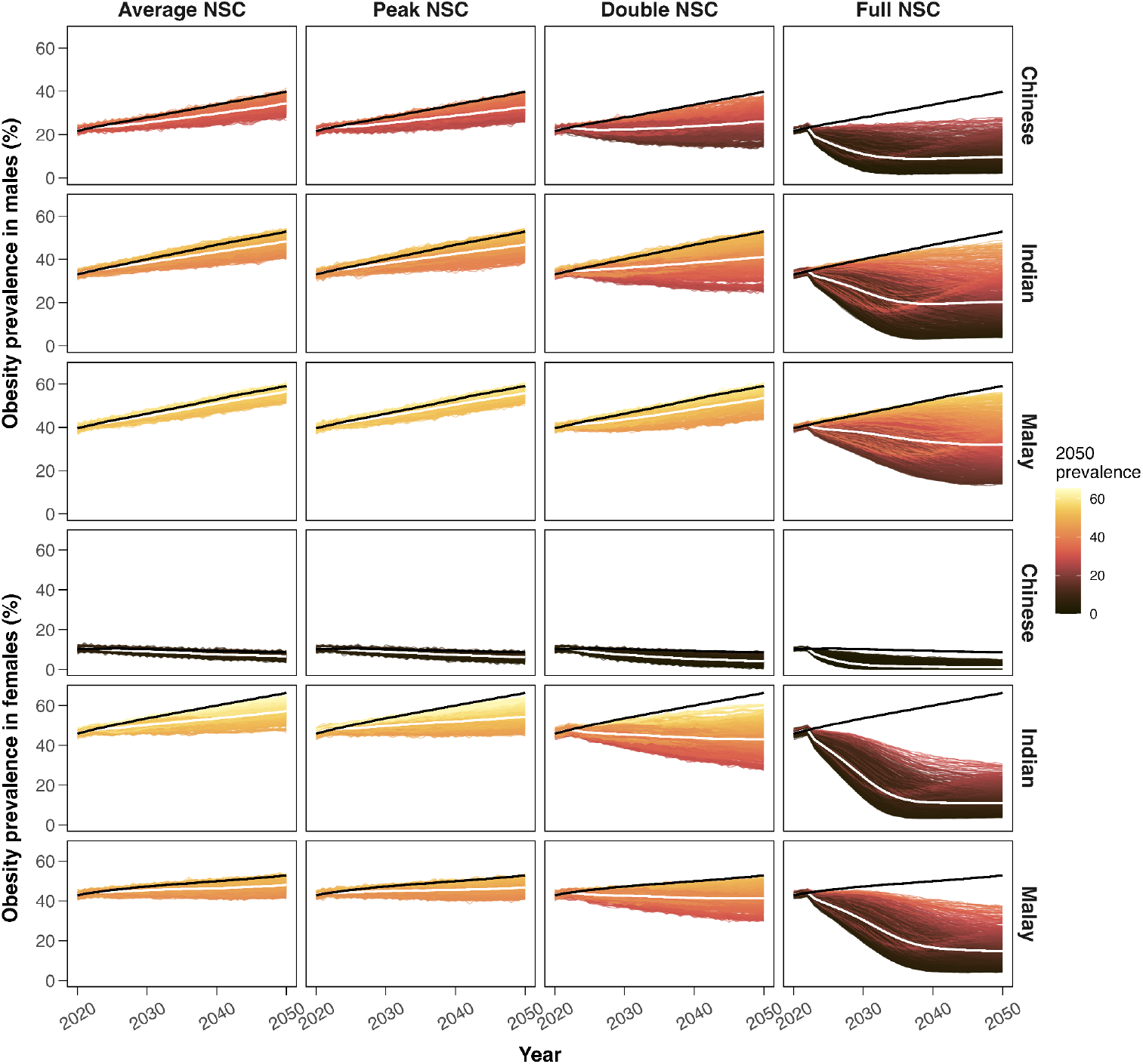
Forecast obesity prevalence rates under four NSC scenarios stratified by sex and ethnicity, which consists of 1000 individual obesity prevalence simulation traces (gradient), median obesity prevalence under the No NSC scenario (black line) and median obesity prevalence under the four NSC scenarios (white line): Average NSC, Peak NSC, Double NSC, and Full NSC.

**Figure 2.**
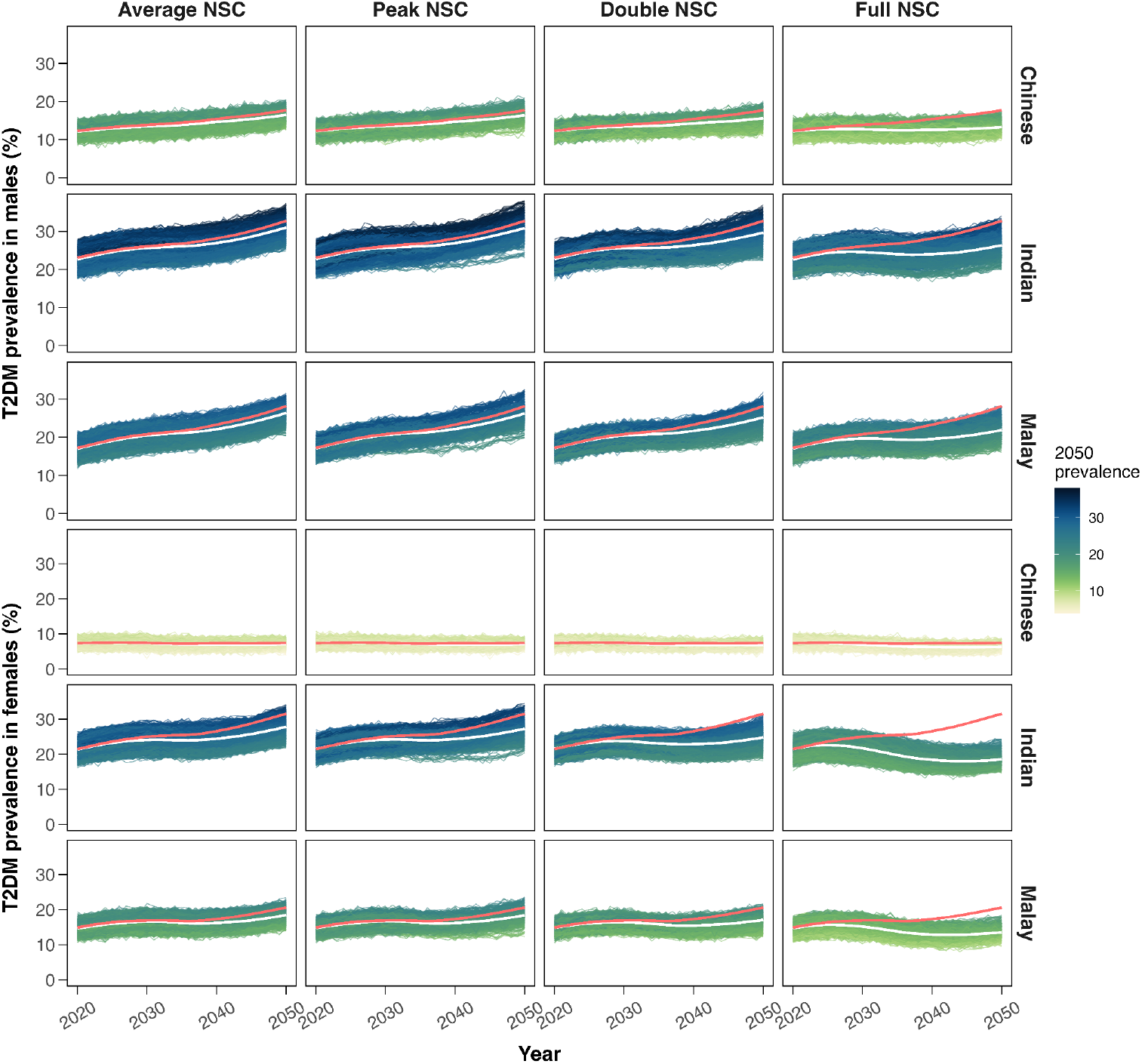
Forecast T2DM prevalence rates under four NSC scenarios stratified by sex and ethnicity, which consists of 1000 individual T2DM prevalence simulation traces (gradient), median T2DM prevalence under the No NSC scenario (red line) and median T2DM prevalence under the four NSC scenarios (white line): Average NSC, Peak NSC, Double NSC, and Full NSC.

Under the Average NSC scenario, Chinese male prevalence is projected to reach 16·5% (13·4%–19·6%), a 6·8% reduction (n=15,083) compared to baseline. Malay and Indian male prevalences are projected to reach 26·4% (22·2%–30·4%) and 31·0% (26·3%–35·8%), with reductions of 6·4% (n=4,862) and 5·8% (n=4,326), respectively. Chinese female prevalence is projected to reach 7·1% (5·3%–8·8%), 4·1% lower (n=3,540) than baseline. Malay and Indian female prevalences are projected to reach 18·4% (14·9%–21·9%) and 27·7% (22·9%– 32·7%), with reductions of 10·7% (n=5,404) and 11·7% (n=6,869), respectively.

Under the Peak NSC scenario, small additional reductions are observed, ranging from 4·1% to 13·3% across the demographic groups. The Double NSC scenario produces more substantial declines. Chinese male prevalence is projected to reach 15·6% (12·5–18·7%), a reduction of 11·9% (n=26,396) from baseline. Malay and Indian male prevalences are projected to reach 25·2% (20·5%–29·4%) and 29·6% (23·9%–35·0%), with reductions of 10·6% (n=8,104) and 9·7% (n=7,286), respectively. Among females, T2DM prevalence stabilises. Chinese female prevalence is projected to reach 6·8% (5·3%–8·3%), an 8·1% reduction (n=7,080) from baseline. Malay and Indian female prevalences are projected to reach 17·0% (13·4%–20·3%) and 24·7% (20·0%–29·3%), reflecting reductions of 17·5% (n=8,843) and 21·6% (n=12,623), respectively.

The Full NSC scenario results in substantial declines across all groups. Chinese male prevalence is projected to drop to 13·3% (10·7%–16·6%), a reduction of 24·9% (n=55,306) from baseline. Malay and Indian male prevalences are projected to reach 21·9% (18·1%– 26·9%) and 26·3% (21·4%–32·2%), reflecting reductions of 22·3% (n=17,019) and 20·0% (n=15,027), respectively. Chinese female prevalence is projected to reach 6·3% (4·9%– 8·0%), 14·9% lower (n=12,980) than baseline. Malay and Indian female prevalences are projected to reach 13·5% (11·0%–16·7%) and 18·5% (15·6%–22·6%), with reductions of 34·5% (n=17,440) and 41·3% (n=24,133), respectively (Figure 2).

## Discussion

This study utilised microsimulation modelling within the DEMOS framework to assess the long-term impact of the NSC on the prevalence of obesity and T2DM in multiethnic Singapore. Based on current recruitment and programme retention rates in the Average NSC scenario, relative to No NSC, total obesity and T2DM prevalence could decrease by 15·8% (n=179,380) and 6·2% (n=36,407), respectively, by 2050. The projected reductions in obesity and T2DM across all demographic groups indicate that physical activity programmes could yield long-term population-level health benefits, which may not be immediately apparent in the programme’s early years, but cumulatively become substantial.

The potential of full population recruitment (Full NSC) demonstrates that a population-wide shift in physical activity levels, driven by sustained behavioural change and infrastructural redesigns that encourage or necessitate active living, could substantially reduce obesity and T2DM prevalence across all demographic groups. Within this scenario, we projected substantial reductions of 74·3% (n=843,541) in obesity and 23·8% (n=139,754) in T2DM relative to No NSC. Such a programme would require a considerable shift in behaviours, leveraging on personal, social, and incentivisation motivations.^28,29^ Desire for weight loss, improved fitness, monetary gain, as well as normalisation of high physical activity levels, are challenging when many Singaporean residents work long hours, face high living costs, and reside in high-calorie accessible environments with considerable escalator and travellator infrastructure.^30–33^

A more attainable goal may be the Double NSC scenario, which models a doubling of both enrolment and retention rates compared to historical averages. Achieving this level of scale-up would necessitate targeted engagement of the remaining 2·9 million unenrolled Singapore residents as well as the 1·17 million prior dropouts (as of October 2023). As physical activity schemes require constant evolution to remain appealing and effective,^34^ rolling surveys on evaluation, engagement drivers, and barriers is necessary to better understand participant and non-participant behaviours. If achieved, based on our microsimulation results, successful deployment of the Double NSC could yield considerable population-wide reductions in obesity and T2DM prevalence by 37·1% (n=421,203) and 11·2% (n=65,766), respectively, by 2050.

Our findings are echoed in another smaller simulation study of 98,000 residents in Los Angeles, USA, which explored the impacts of 14 different intervention types on T2DM risk. Individuals who met daily moderate to vigorous physical activity requirements were estimated to have their risk ratio of developing T2DM reduced to 0·847 (95% CI 0·730– 0·951), supporting the promotion of local physical activity schemes to reduce chronic disease burden risk.^35^ The implementation of such schemes was furthermore greatly improved when using digital approaches to set goals (β = 0·89, p = 0·001), grade performance (β = 0·87, p = 0·008), and provide group incentives (β = 2·37, p < 0·001).^28,29^

With rising costs of T2DM treatment and associated chronic diseases, physical activity interventions could be especially cost-effective if designed to be self-motivated with support from peers and general practitioners, as is the focus of the NSC and other schemes.^36,37^ At the population level in Australia, a 5% reduction in new cases of obesity was estimated to increase productivity-adjusted life years by 1229 over 10 years, adding the economic equivalent of US$165 million to the GDP.^36^ Through simple extrapolation, this could be US$4·3 billion for Southeast Asia. Reductions in obesity could thus bring cost-savings to many healthcare systems under considerable strain.^37^ This could be very advantageous with rising outdoor temperatures, which is projected to increase the risks of negative outcomes among T2DM sufferers due to the projected increasing number of heat days, where the promotion of indoor activities through schemes such as the NSC, and further investments in indoor exercise facilities, could assist in alleviating this burden.^38,39^

The downstream impact of the NSC is expected to also impact mortality and other conditions, or diseases. Mortality in 10·6 million individuals has been found to increase log-linearly throughout the overweight and obese BMI ranges, with a 1·39 (95% CI 1·34–1·44) increase in hazards ratio for every five unit BMI increase starting from 25 kg/m^2^.^40^ With a 10% weight loss in obese individuals, averted years spent with hypertension and T2DM were estimated at 1·2–2·9 and 0·5–1·7 years respectively, and the number of coronary heart disease cases is estimated to decline by 13 to 38 cases per 1000 persons.^41^ With a 2-5% weight loss in obese individuals, the odds of having clinical improvements in systolic blood pressure (SBP) (odds ratio 1·24 [95% CI 1·02–1·50]), glucose levels (1·75 [1·40–2·19]), HbA1c (1·80 [1·44– 2·24]), and triglycerides (1·46 [1·14–1·87]) were substantial.^42^ Utilising programmes such as the NSC could thus assist in alleviating multiple NCD burdens by addressing increasing obesity rates.

Furthermore, the NSC could be combined with dietary interventions, pharmacotherapy, and surgery. Caloric restrictions through portion control and diet composition can result in lower T2DM and obesity risks, complementing physical activity programmes.^43^ The prescription of weight-loss medication such as Wegovy or Orlistat, to people with obesity, or bariatric surgery, could also be run in tandem under clinical guidance.^44–46^ Meeting required daily physical activity levels enables patients to sustain the effects of such dietary, medication, and surgical interventions as part of the national promotion of physical activity, normalising healthier lifestyles.

Our study has several limitations however. We modelled changes in BMI due to physical activity using demographic variables, current BMI, and physical activity metrics, thus averaging over other factors such as participants’ resting metabolic rate and changes in diet.^47^ Additionally, we modelled physical activity measures (basesteps and deltasteps) using demographic variables and BMI, while averaging over other factors such as chronic conditions and diseases, depression, prior exercise adherence, current year, and environmental determinants.^48–50^ Due to the limited availability of such data we omitted these variables from our study, but if such data become available, it should be incorporated into the existing model to achieve higher accuracy in estimating individual physical activity levels and the related health impact. Our study also leaves out some of the demographic groups that were not represented in the NSC, such as individuals with physical or mental disabilities who are unable to participate in such programmes. We should be cautious about extrapolating the results to such populations. Additionally, BMI might not be a wholly representative measure of health and determinant of health outcomes, where body fat measures could be explored.^51^ Lastly, adherence to physical activity programmes and weight gain could vary considerably between individuals, and although our model captures the population level, variations are expected within smaller subgroups, which could exhibit unknown trends over time.

## Conclusions

Our microsimulation model of 7·9 million simulated individuals explored the impacts of a national physical activity programme on obesity and T2DM prevalences. The current programme estimated that relative to No NSC, total obesity and T2DM prevalence could decrease by 15·8% (n=179,380) and 6·2% (n=36,407), respectively, by 2050. We found that scaling this up to the Double NSC enrolment and retention levels could reduce obesity and T2DM prevalence by 37·1% (n=421,203) and 11·2% (n=65,766), respectively, by 2050. The NSC or similar programmes could thus be utilised with continuing evolution, to assist in alleviating rising chronic disease burdens.

## Data Availability

The datasets generated and/or analysed during the current study are not publicly available due to the participants' privacy but aggregates are available from the corresponding author upon reasonable request.

## Contributions

BN, HS, VFZ, YH, XZ, and BLD developed simulation model components. BN and BLD designed the study. BN conducted data processing and data analysis. BN and BLD wrote the original manuscript draft, and all authors contributed to critically reviewing and revising the manuscript. All authors read and approved the final manuscript.

## Declaration of Interests

The authors declare that they have no competing interests.

## Acknowledgements

We thank all the research staff and investigators who contributed to this project. This study was supported by a grant from the Health Promotion Board, Singapore. The funders had no role in the conceptualization, design, analysis, decision to publish or preparation of the manuscript.

## Author Information

Saw Swee Hock School of Public Health, National University of Singapore and National University Health System, Singapore Bekzod Normatov, Haolong Song, Vong Fee Zheng, Yichen He, Xinyu Zhang, Nigel WH Lim, Hafiz BM Aziz, Shihui Jin, Borame L Dickens

## List of abbreviations

BMI: Body mass index
COVID-19: Coronavirus Disease of 2019
DEMOS: Demographic epidemiological model of Singapore
GDP: Gross Domestic Product
HPB: Health Promotion Board Hb
A1c: Glycated hemoglobin
MCMC: Markov Chain Monte Carlo
MLE: Maximum Likelihood Estimator
NSC: National Steps Challenge
NSCAO: National Steps Challenge Always-On
PA: Physical activity
RCT: Randomized control trial
T2DM: Type 2 diabetes mellitus
USA: United States of America

## Notes

### Competing Interest Statement

The authors have declared no competing interest.

### Author Declarations

The Department Ethics Review Committee of the Saw Swee Hock School of Public Health waived ethical approval for this work.

