## Supplementary Information for "The role of physical activity programmes in mitigating obesity and type 2 diabetes in Singapore using simulation-based forecasts until 2050"

### Supplementary Data

**Supplementary Information 1. Demographic Epidemiological Model of Singapore (DEMOS) specification.** **2**
Sample size 2
BMI model 2
Fertility Model 5
Migration model 6
Diabetes model 7
Physical activity model 8
**Supplementary Information 2. NSC participation models.** **23**
First time participant model 24
Continuing participant model 25
**References** **26**

### Supplementary Information 1. Demographic Epidemiological Model of Singapore (DEMOS) specification.

#### Sample size

The total sample size of 7,956,003 individuals covered Singaporean adult residents aged 18-74 from 1990 to 2050. We used the Demographic Epidemiological Model of Singapore (DEMOS) to generate a synthetic population representing each resident and citizen of Singapore from 1990 to 2050.

We validated our sample size in the microsimulation by comparing the total simulated number of residents aged 20 to 74 from 1990 to 2024 and the number of annual residents provided by the Singapore Department of Statistics as seen in Figure S1.[1] Our estimates, while slightly deviating, mostly align and the mean average percentage error is less than 1% (MAPE = 0.962%).

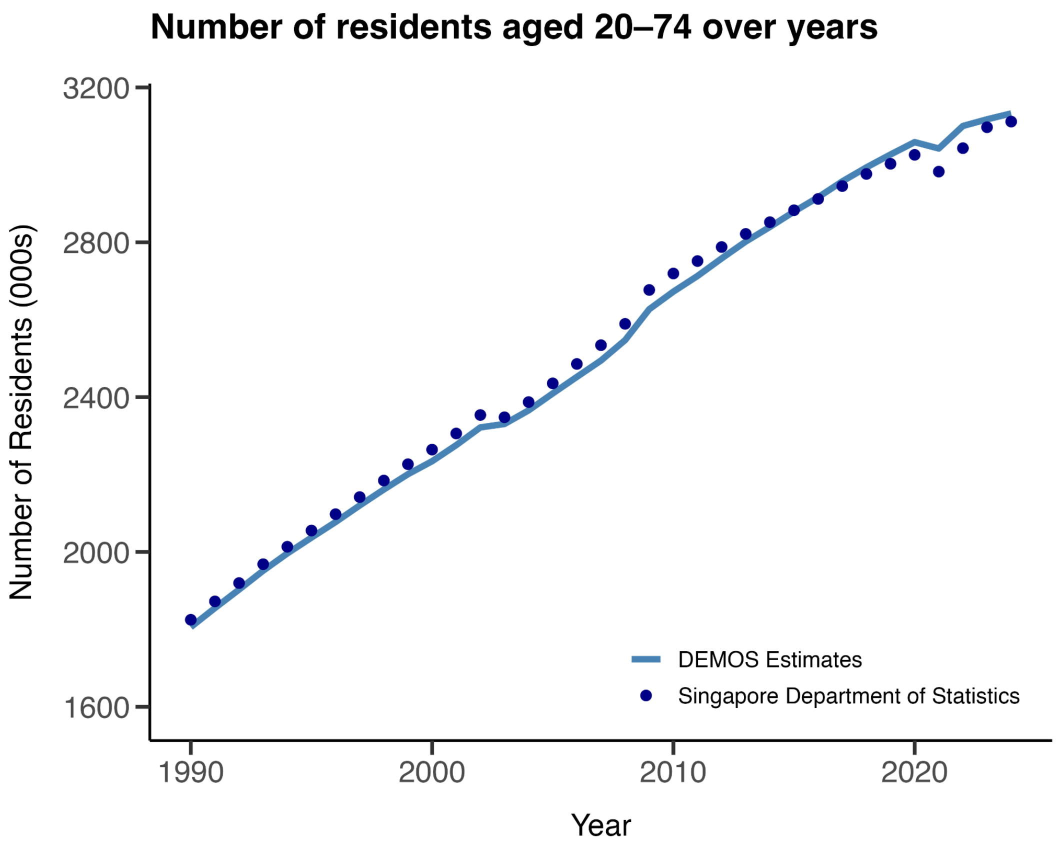


**Figure S1.** Comparison between the number of total adult residents aged 20-74 in Singapore as reported by the Singapore Department of Statistics and DEMOS estimates from 1990 to 2024.

#### BMI model

For BMI, we created a hierarchical model to track individual changes in BMI across adulthood based on sex and ethnicity. We simulated an individual's BMI trajectory as normal fluctuations around piecewise connected lines. BMI values at age 18 and following gradients vary by individual but follow group-specific distributions (Figure S2). The hyperparameters of these distributions were determined using BMI measurements from the Singapore Prospective Study programme (1992-2005) and validated against national population health surveys (2004, 2010) that provided BMI category proportions (underweight, normal weight, overweight, and obese) across age, ethnicity and sex groups.

Individual BMI trajectories were simulated using a stochastic, piecewise polynomial model parameterised by demographic group, birth cohort, and random variation. Each individual belonged to one of eight demographic groups indexed by an integer variable corresponding to: 0 = male Chinese, 1 = female Chinese, 2 = male Malay, 3 = female Malay, 4 = male Indian, 5 = female Indian, 6 = male Other, and 7 = female Other.

For each individual, a set of hyperparameters ($M_1$,$M_2$,$M_3$,$M_4$) was generated as a linear combination of demographic means and random effects. Then, log-BMI values were computed and fixed at ages 18 ($y_0$), 35 ($y_1$ ), 55 ($y_2$) and 75 ($y_3$) dependant on a year of birth ($y_b$) and a demographic group ($b_i$):

$$y_0 = M_2 + b_i(y_b - 1950) + M_1(35 - 18);$$

$$y_1 = M_2 + b_i(y_b - 1950);$$

$$y_2 = M_2 + b_i(y_b - 1950) + M_3(55 - 35);$$

$$y_4 = M_2 + b_i(y_b - 1950) + M_3(35 - 18) + M_4(75 - 55)$$

Between these fixed points, log-BMI at age *a* was estimated using piecewise cubic polynomials:

$$log(BMI(a)) = y_0 + b_0(a - 18) + d_0(a - 18)^3 \; for \; 18 \le a < 35$$

$$log(BMI(a)) = y_1 + b_1(a - 35) + \frac{m_1}{2}(a - 35)^2 + d_1(a - 35)^3 \; for \; 35 \le a < 55$$

$$log(BMI(a)) = y_2 + b_2(a - 55) + \frac{m_2}{2}(a - 55)^2 + d_2(a - 55)^3 \; for \; 55 \le a < 80$$

$$log(BMI(a)) = y_2 + b_2(25) + \frac{m_2}{2}(25)^2 + d_2(25)^3 \; for \; a \ge 80$$

where the coefficients $b_0$, $b_1$, $b_2$, $m_1$, $m_2$, $d_0$, $d_1$, $d_2$ were determined algebraically to ensure continuity and smoothness at the boundaries of each segment. For every age, the individual's log-BMI was modified by adding a normally distributed random error with standard deviation σ (specific to each demographic group, from the parameter table) and then exponentiated to yield BMI in $kg/m^2$:

$$BMI(a) = exp(N(log(BMI(a)), \sigma^2)).$$

Finally, to ensure plausibility of generated BMI values, simulations were constrained such that log-BMI values at the anchor ages and at age 80 remained within the interval [2.6,4.2], corresponding to the plausible BMI range [13.5, 66.7]. Figure S2 shows a sample of simulated BMI trajectories of the representative adult population from 1990 to 2050 stratified by three major ethnic groups and two sexes.

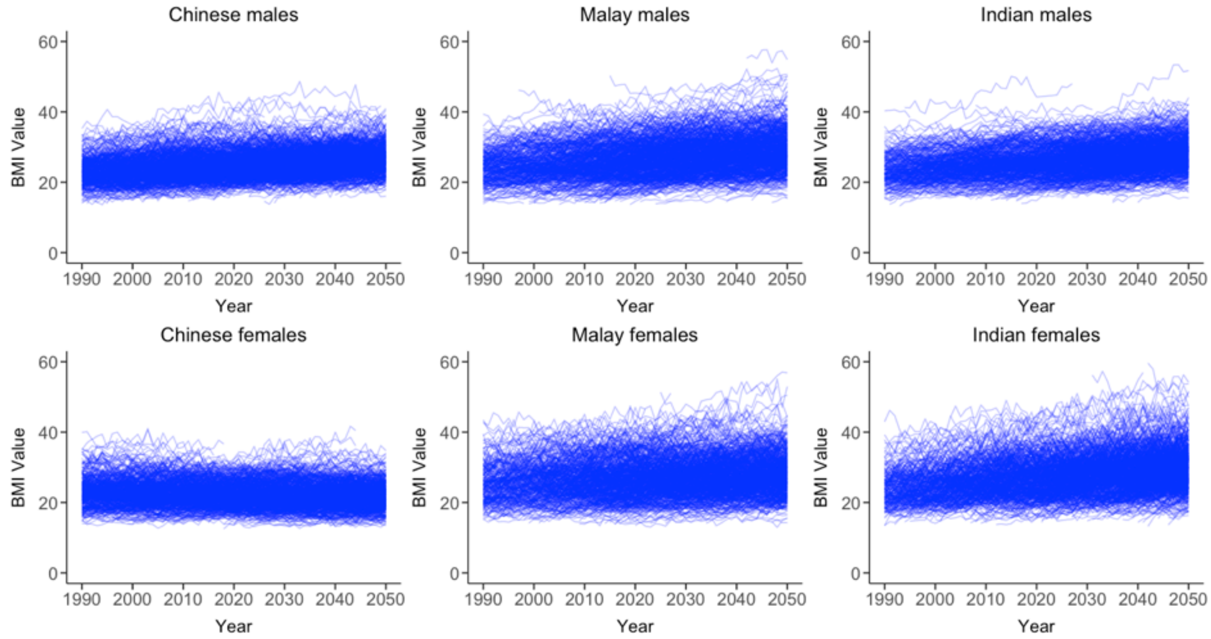


**Figure S2.** Individual simulated BMI trajectories of Singaporean adults (ages 18-74) by three major ethnicities and sex (n=1,000).

We further validated the BMI trajectories of our microsimulation model by computing population-wide obesity prevalence as defined by BMI ≥ 30 kg/m² and comparing DEMOS estimates with the National Population Health Survey data, which can be seen on Figure S3.[2,3] All NPHS estimates fall within 95% simulation interval of the DEMOS runs.

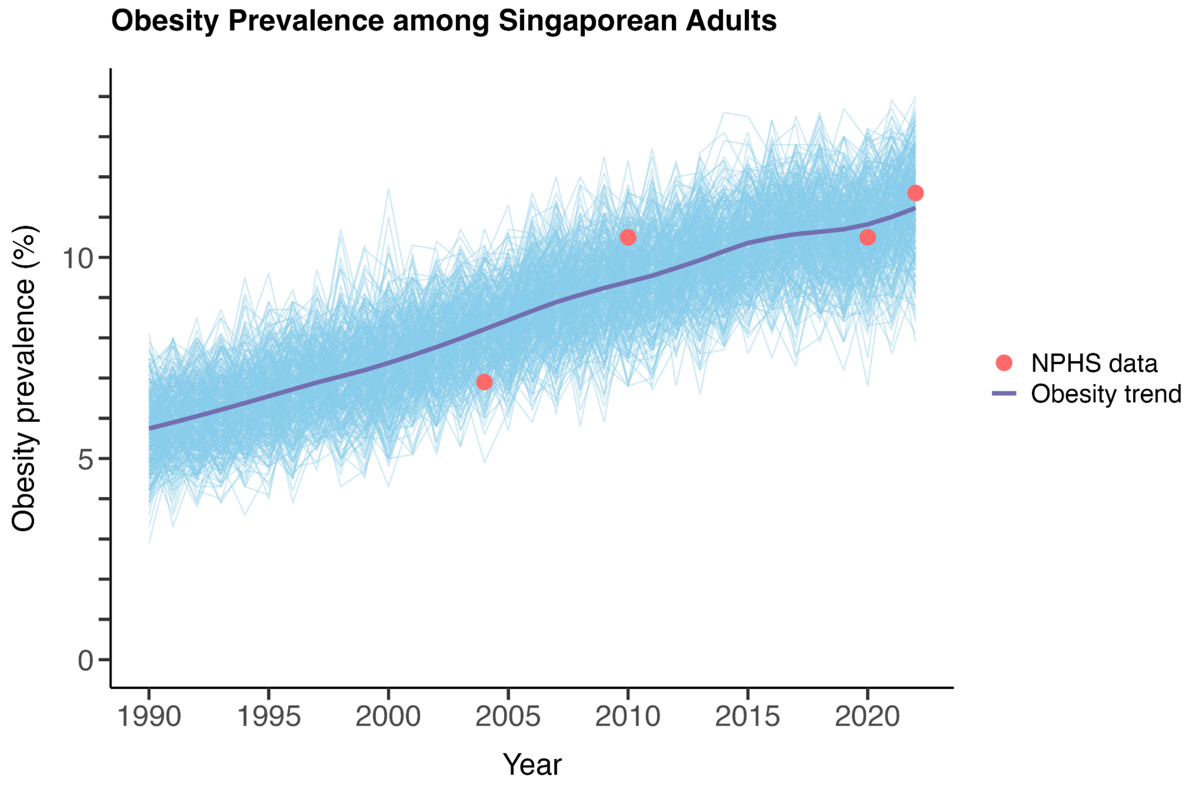


**Figure S3.** 1000 simulated obesity prevalence (BMI ≥ 30 kg/m²) trajectories for all adults with National Population Health Survey (NPHS) data overlaid. Median T2DM trend is in purple, individual trajectories are in light blue and NPHS data points are in red.

### Fertility Model

In Singapore, there are three major ethnic groups: Chinese (74%), Malays (13%), and Indians (9%), and they exhibited distinct fertility patterns. To predict total fertility rates of these distinct groups we utilize a fertility model which has been described elsewhere.[4] For populations not included in these three ethnic groups, we inferred their fertility rates based on those of the Malays, given the limited sample size and the similarity in observed values.

The datasets used for modelling comprised annual fertility rates for each ethnic group during the 'low fertility phase'—a period starting when two consecutive decreases in fertility rates resulted in levels dropping below the 2.1 threshold.[5] This phase extended from 1975 to 2022 for Chinese and Indians, and from 2003 to 2022 for Malays. Bayesian Structural Time Series (BSTS) models were applied to generate long-term forecasts. To better reflect the slowing decline during this phase, a logarithmic transformation was used.

Let $F_t$ denote the logarithm of fertility rate in year $t$. For each ethnic group, the BSTS model is defined as $F_t = \mu_t + s_t + \epsilon_t$, $\epsilon_t \sim N(I, \sigma_\mu)$, where $\mu_t$ represents the trend, $s_t$ represents the seasonality effect and $\epsilon_t$ represents the residual. For the Chinese and Indian ethnic groups, a semi-local linear trend model was used, which can be represented as

$$\mu_{t+1} = \mu_t + \delta_t + \eta_{0t},$$

$$\delta_{t+1} = D + \phi(\delta_t - D) + \eta_t, \ \eta_{1t} \sim N(I, \sigma_{\delta t}),$$

where $D$ is the long term gradient of the trend component, towards which $\delta_t$ will eventually revert, $\phi$ determines the memory in autoregressive (AR) deviations from the long term trend, and both $\eta_{0t}$ and $\eta_t$ are random noise components. For the Malay ethnic group, due to the relative simple pattern of the time series an AR(1) model was used

$$\mu_{t+1} = \phi\mu_t + \eta_t,$$

where $|\phi| < 1$ and $\eta_t$ is random noise. Finally, the seasonality parameter was non-zero and time-dependent only in the model for the Chinese ethnic group, where the fertility rates exhibited a 12-year cyclic pattern.

For each ethnic group, we split the time series into training, testing and validation sets and simulated 5000 MCMC draws from the model. We used mean values of these draws as point estimates of fertility rates. In terms of performance, the models demonstrated high prediction accuracy across the validation sets as can be seen in Table S1.

**Table S1.** Model performance, measured by main absolute percentage error (MAPE) for the three BSTS models.

| | **Training set** (year) | **Validation set** (year) | **$MAPE_{training}$** (%) | **$MAPE_{validation}$** (%) |
|---|---|---|---|---|
| **Chinese** | 1975–2010 | 2011–2022 | 4.76 | 3.71 |
| **Malays** | 2003–2017 | 2018–2022 | 5.91 | 4.69 |
| **Indians** | 1975–2010 | 2011–2022 | 3.33 | 4.77 |

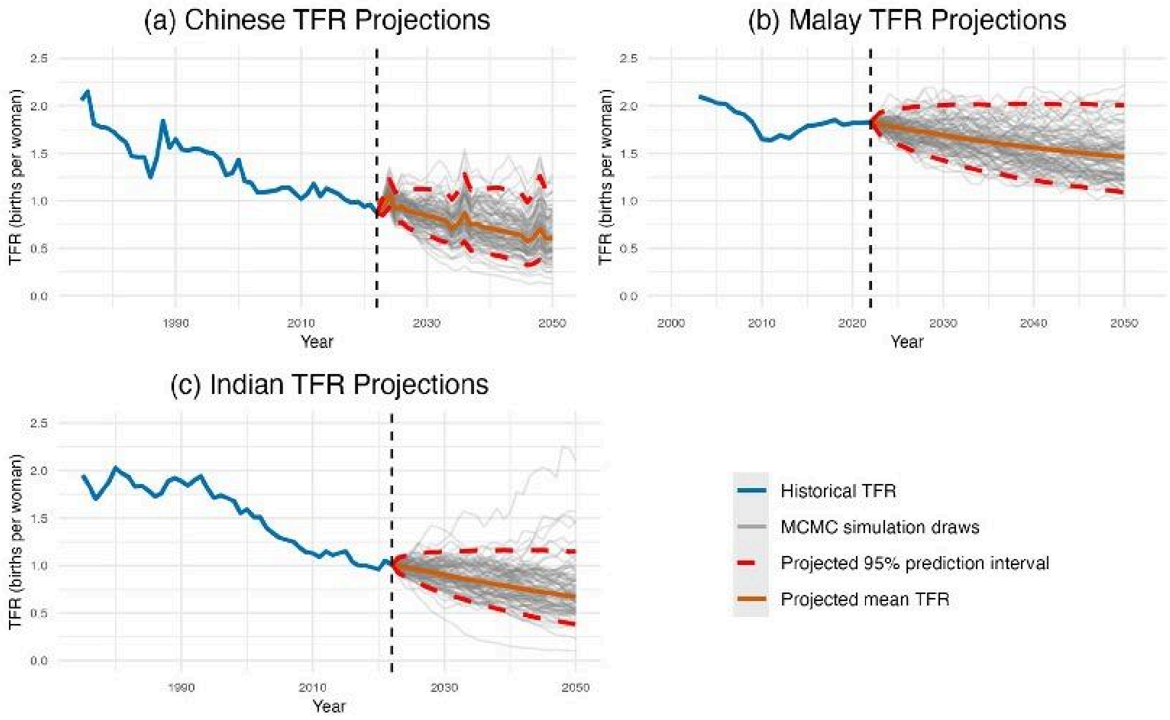


**Figure S4.** Projected total fertility rates for Chinese (a), Malays (b), and Indians (c) from 2023 to 2050, with grey lines representing a sample of 100 MCMC simulation draws.

**Mortality model**

We used age-specific averaged mortality rate data from 2016 to 2019 for mortality projections as rates have remained stable during this period and beyond as seen below.[6]

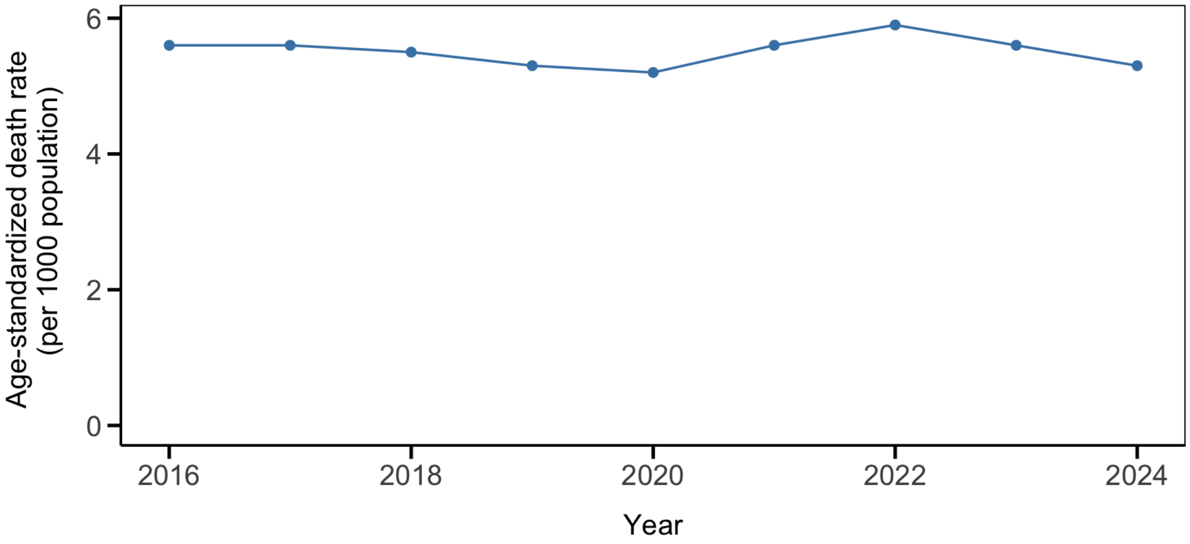


**Figure S5.** Age-standardized mortality rates in Singapore (per 1000 residents) from 2016 to 2024.

**Migration model**

We estimated yearly migration rates from 1990 to 2020 using fertility and mortality rates published by the Singapore Department of Statistics for the same time period as seen in the figure below. For future projections, we assumed the migration rates will remain similar with small variance starting from 2010 onwards. Hence, we estimated the mean and variance of migration rates based on data from 2010 to 2020, and modeled future migration as a stochastic process using these parameters.

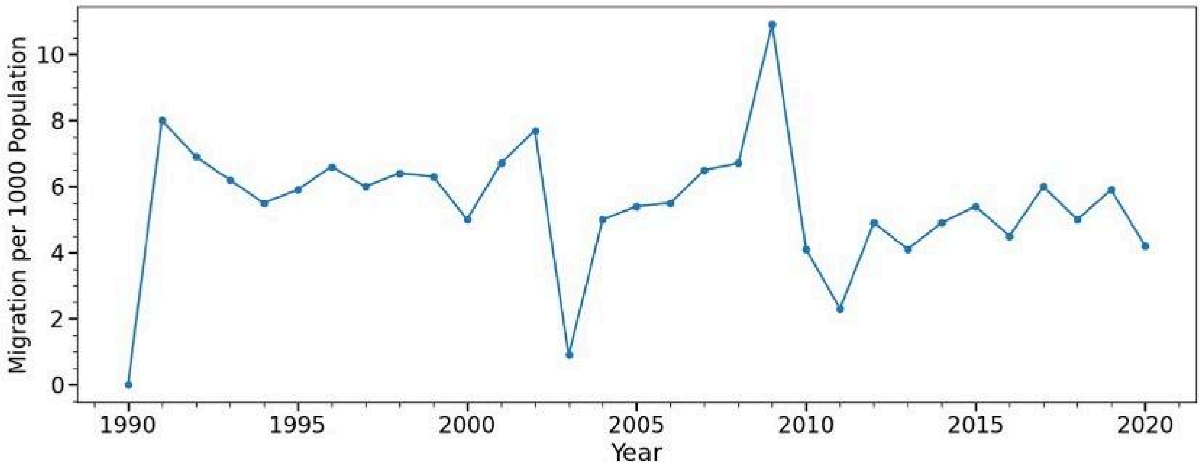


**Figure S6.** Singapore migration trends (number of migrants per 1000 resident population) for 1990-2020.

**Diabetes model**

Type 2 diabetes mellitus (T2DM) prevalence data came from the Singapore Prospective Study programme containing T2DM status at two of three time points (1992, 1998, 2005). We developed a logistic model to estimate the probability of developing T2DM given a non-diabetic state. For every individual we generated a BMI trajectory, which served as a predictor together with age, gender and ethnicity. Within the simulation, the probability of developing T2DM was calculated annually as a logistic function using demographic data and BMI of each individual [23,24]:

$$P(Diabetes\ at\ age\ a) = 1 - \frac{1}{1+exp(\theta_{index}+\delta a+\beta BMI(a))},$$

where $a$ is the individual's age, $BMI(a)$ is the individual's BMI at age $a$, $\theta_{index}$ is a baseline log-odds parameter specific to the individual's demographic group (with groups defined by gender and ethnicity, $\delta$ and $\beta$ are coefficients representing the effect of age and BMI, respectively, derived as point estimates of the 5000 MCMC draws. For each simulated year, a random number between 0 and 1 is drawn from a uniform distribution. If this random number is less than the calculated probability,$P(Diabetes\ at\ age\ a)$, the individual is considered to have developed diabetes in that year. The first year in which this event occurs is recorded as the year of diabetes onset for the individual. If the threshold is not met, the individual does not develop diabetes in that year. Using simulated year of diabetes onset, we computed T2DM prevalence in the population from 1990 to 2024 and validated the results on the data provided by the National Population Health Survey (NPHS) in Singapore. See Figure S7 for simulated T2DM prevalence.

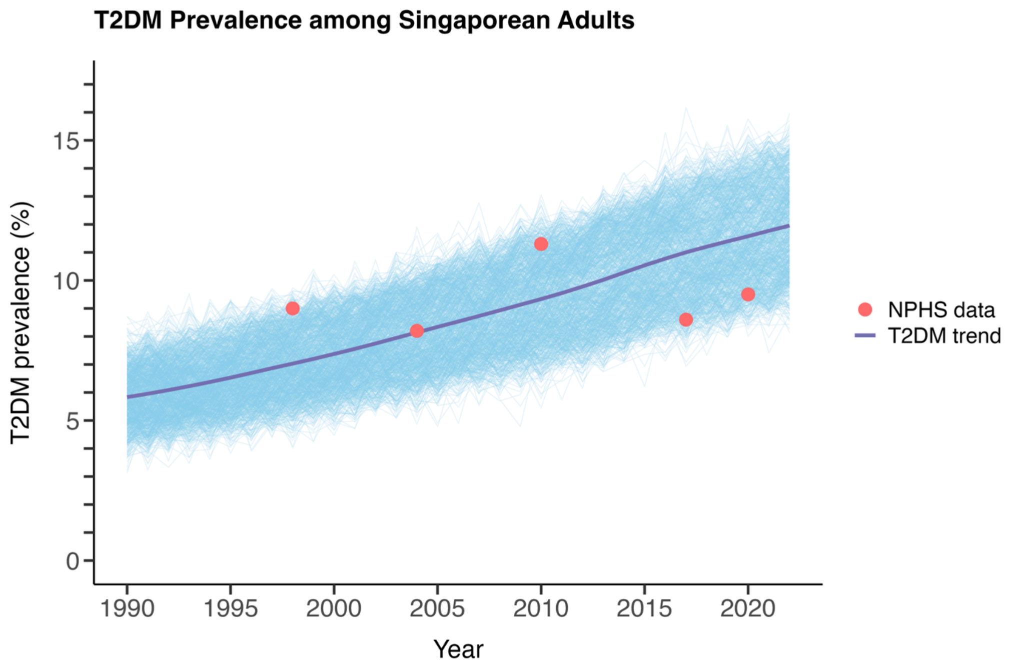


**Figure S7**. 1000 simulated Type 2 diabetes mellitus (T2DM) prevalence trajectories for all adults with National Population Health Survey (NPHS) data overlaid. Median T2DM trend is in purple, individual trajectories are in light blue and NPHS data points are in red.

**Physical activity model**

We used average daily stepcount as a proxy for physical activity levels in the population. We utilized the National Steps Challenge (NSC) datasets which included NSC participants' daily step count data to model physical activity patterns across demographic groups in Singapore. Each individual enrolled into the NSC had baseline step count as well as change in step count during the intervention phase. Baseline step count for each NSC participant was obtained by averaging the participants' daily average non-zero step count data during the pre-intervention period (11 weeks before the start of the incentive period in NSC3, NSC4 and NSC5). We included only those baseline step count records that are equal to or greater than 1000 steps, leaving a total of 120,575 participants. Individuals' baseline step count values differed by sex, ethnicity, age and BMI group. We modelled baseline step count as gamma fluctuations with a hyper parametric shape and scale common to all the individuals of a specific demographic strata, with 72 demographic strata in total as can be seen in Figure S8.This is comprised of two sexes (Male, Female), four ethnic groups (Chinese, Malay, Indian, Other), three age groups (18-35, 35-50, 50+) and three BMI risk groups (Low, Medium. High). Point estimates of the shape and scale parameters for each demographic strata were obtained using the Maximum Likelihood Estimator (MLE) as seen in Table S2.

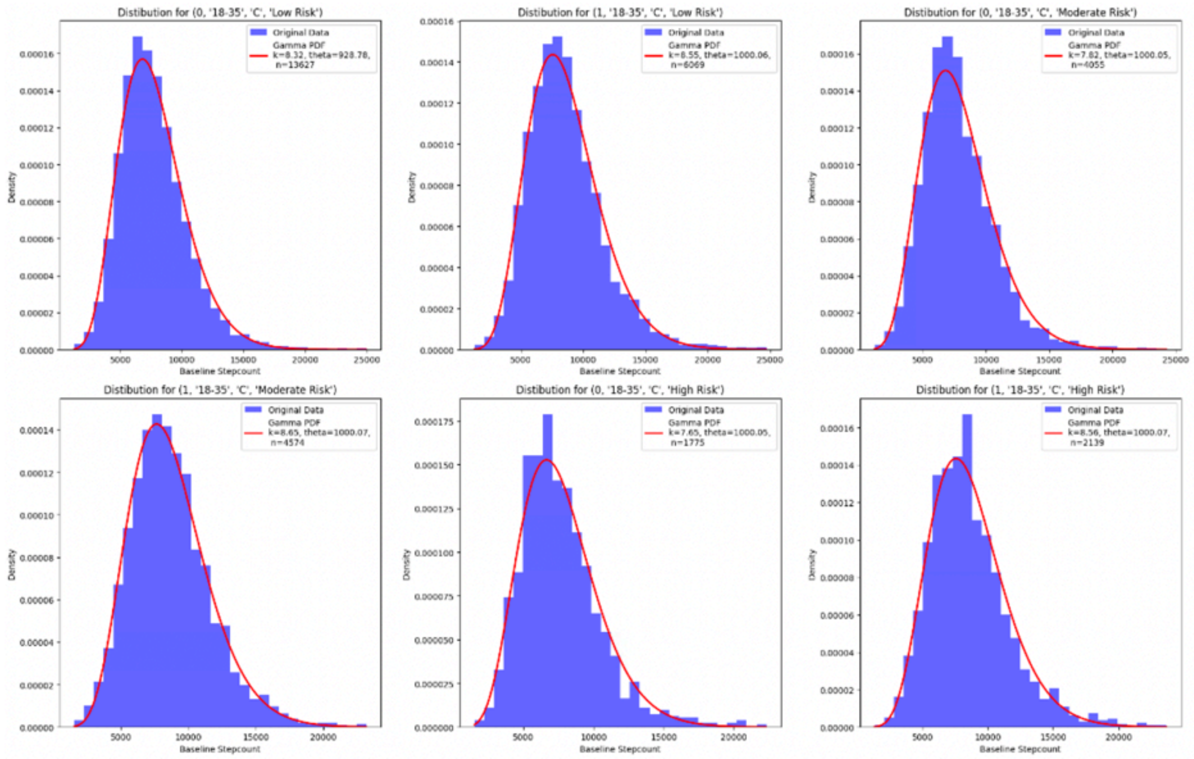


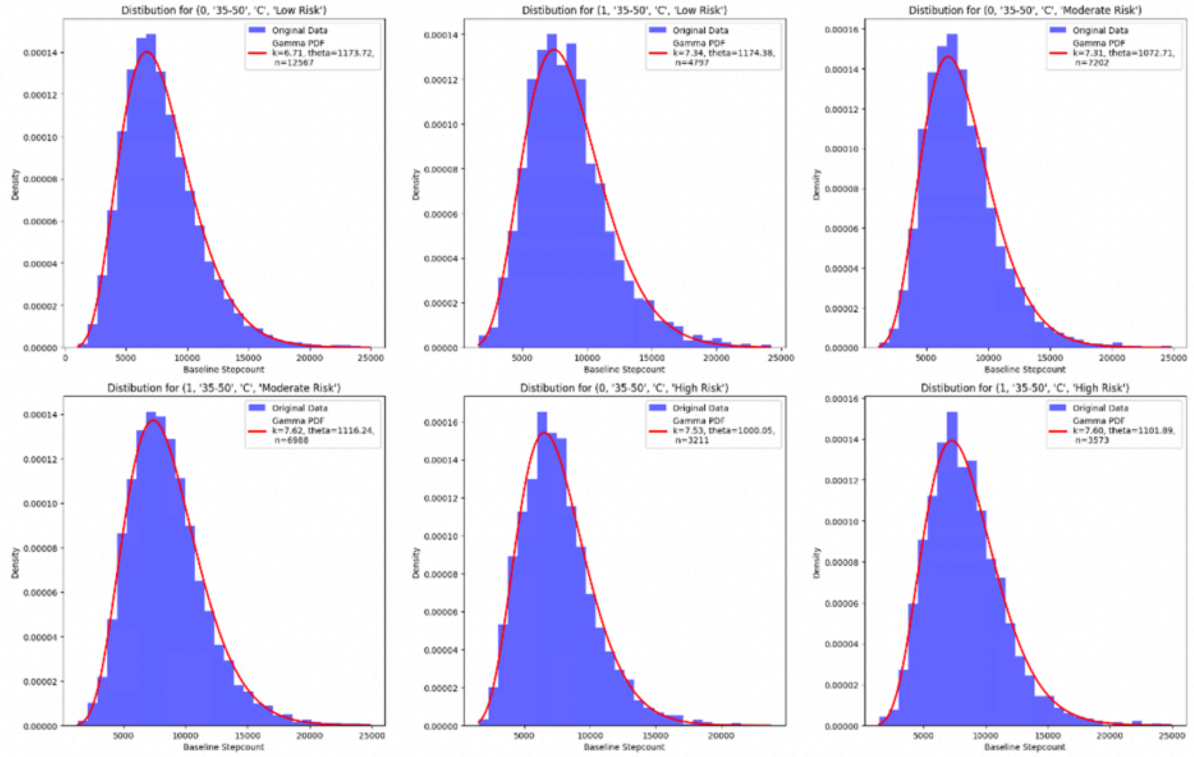

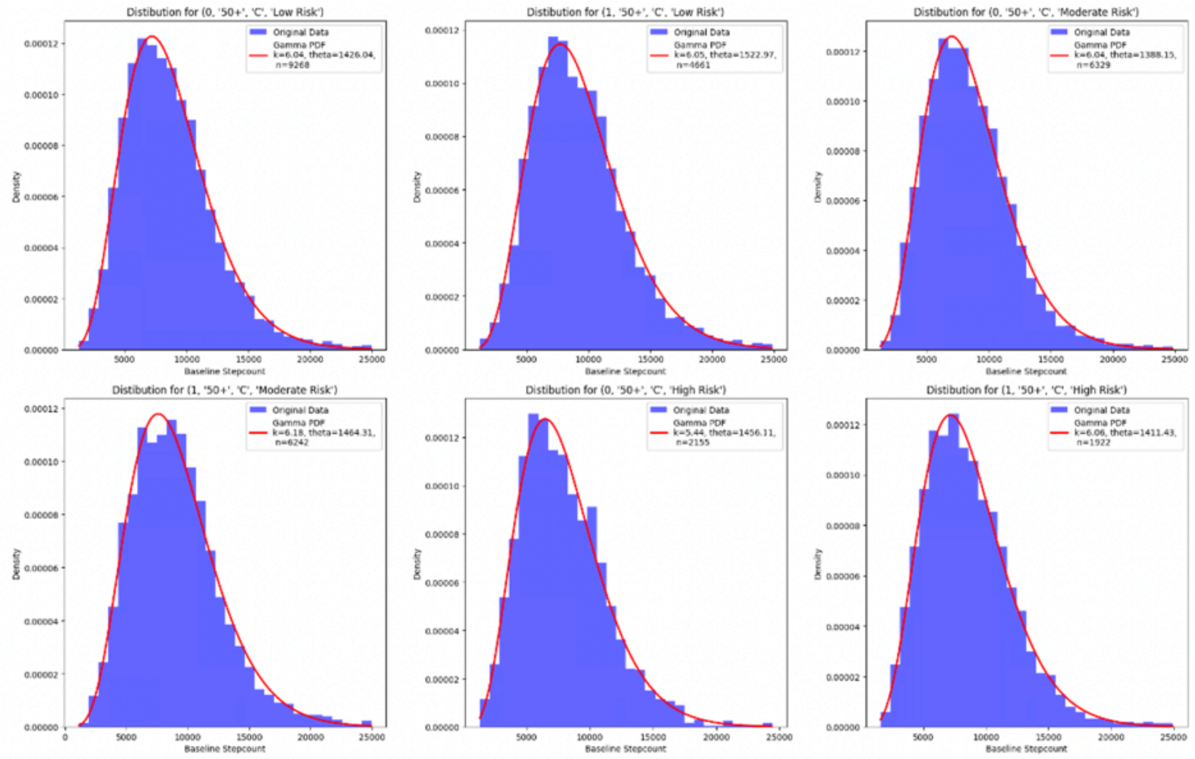
Distibution for (0, '50+', 'C', 'Low Risk')
Original Data
Gamma PDF k=6.04, theta=1426.04, n=9268
Distibution for (1, '50+', 'C', 'Low Risk')
Original Data
Gamma PDF k=6.05, theta=1522.97, n=4661
Distibution for (0, '50+', 'C', 'Moderate Risk')
Original Data
Gamma PDF k=6.04, theta=1388.15, n=6329
Distibution for (1, '50+', 'C', 'Moderate Risk')
Original Data
Gamma PDF k=6.18, theta=1464.31, n=6242
Distibution for (0, '50+', 'C', 'High Risk')
Original Data
Gamma PDF k=5.44, theta=1456.11, n=2155
Distibution for (1, '50+', 'C', 'High Risk')
Original Data
Gamma PDF k=6.06, theta=1411.43, n=1922
Density
Baseline Stepcount

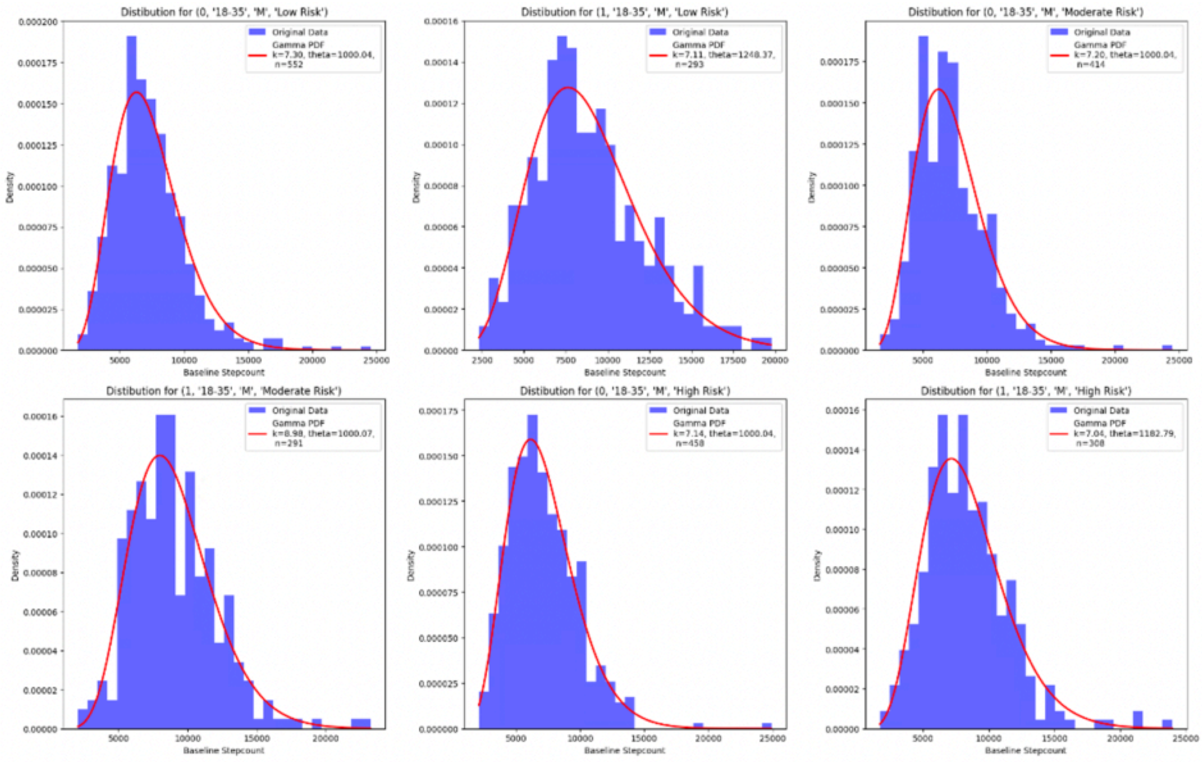
Distibution for (0, '18-35', 'M', 'Low Risk')
Original Data
Gamma PDF k=7.30, theta=1000.04, n=552
Distibution for (1, '18-35', 'M', 'Low Risk')
Original Data
Gamma PDF k=7.11, theta=1248.37, n=293
Distibution for (0, '18-35', 'M', 'Moderate Risk')
Original Data
Gamma PDF k=7.20, theta=1000.04, n=414
Distibution for (1, '18-35', 'M', 'Moderate Risk')
Original Data
Gamma PDF k=8.98, theta=1000.07, n=291
Distibution for (0, '18-35', 'M', 'High Risk')
Original Data
Gamma PDF k=7.14, theta=1000.04, n=458
Distibution for (1, '18-35', 'M', 'High Risk')
Original Data
Gamma PDF k=7.04, theta=1182.79, n=308
Density
Baseline Stepcount

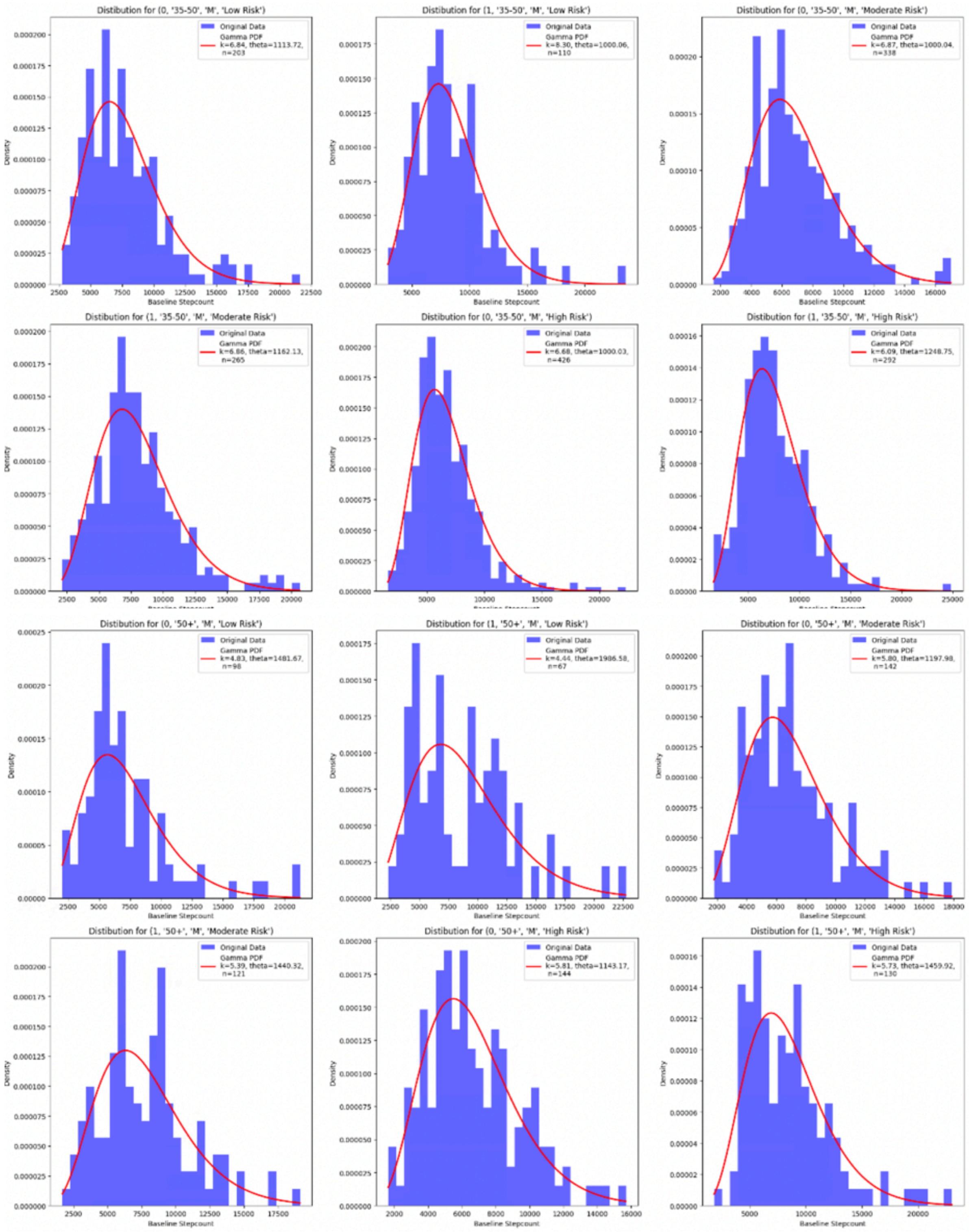
Distibution for (0, '35-50', 'M', 'Low Risk')
Original Data
Gamma PDF k=6.84, theta=1113.72, n=203
Baseline Stepcount
Distibution for (1, '35-50', 'M', 'Low Risk')
Original Data
Gamma PDF k=8.30, theta=1000.06, n=110
Baseline Stepcount
Distibution for (0, '35-50', 'M', 'Moderate Risk')
Original Data
Gamma PDF k=6.87, theta=1000.04, n=338
Baseline Stepcount
Distibution for (1, '35-50', 'M', 'Moderate Risk')
Original Data
Gamma PDF k=6.86, theta=1162.13, n=265
Distibution for (0, '35-50', 'M', 'High Risk')
Original Data
Gamma PDF k=6.68, theta=1000.03, n=426
Distibution for (1, '35-50', 'M', 'High Risk')
Original Data
Gamma PDF k=6.09, theta=1248.75, n=292
Distibution for (0, '50+', 'M', 'Low Risk')
Original Data
Gamma PDF k=4.83, theta=1481.67, n=98
Baseline Stepcount
Distibution for (1, '50+', 'M', 'Low Risk')
Original Data
Gamma PDF k=4.44, theta=1986.58, n=67
Baseline Stepcount
Distibution for (0, '50+', 'M', 'Moderate Risk')
Original Data
Gamma PDF k=5.80, theta=1197.98, n=142
Baseline Stepcount
Distibution for (1, '50+', 'M', 'Moderate Risk')
Original Data
Gamma PDF k=5.39, theta=1440.32, n=121
Baseline Stepcount
Distibution for (0, '50+', 'M', 'High Risk')
Original Data
Gamma PDF k=5.81, theta=1143.17, n=144
Baseline Stepcount
Distibution for (1, '50+', 'M', 'High Risk')
Original Data
Gamma PDF k=5.73, theta=1459.92, n=130
Baseline Stepcount

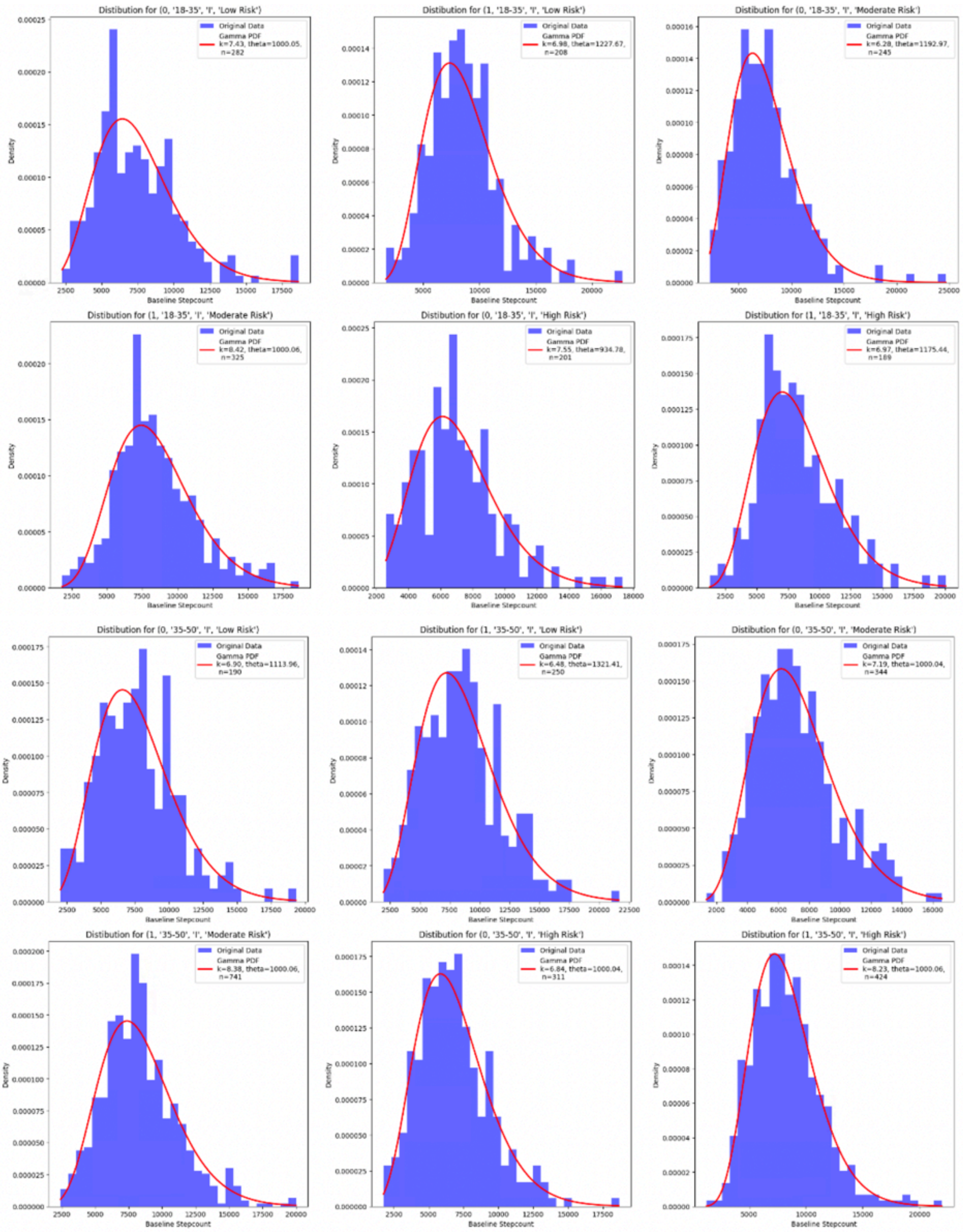
Distibution for (0, '18-35', 'I', 'Low Risk')
Original Data
Gamma PDF k=7.43, theta=1000.05, n=282
Distibution for (1, '18-35', 'I', 'Low Risk')
Original Data
Gamma PDF k=6.98, theta=1227.67, n=208
Distibution for (0, '18-35', 'I', 'Moderate Risk')
Original Data
Gamma PDF k=6.28, theta=1192.97, n=245
Distibution for (1, '18-35', 'I', 'Moderate Risk')
Original Data
Gamma PDF k=8.42, theta=1000.06, n=325
Distibution for (0, '18-35', 'I', 'High Risk')
Original Data
Gamma PDF k=7.55, theta=934.78, n=201
Distibution for (1, '18-35', 'I', 'High Risk')
Original Data
Gamma PDF k=6.97, theta=1175.44, n=189
Distibution for (0, '35-50', 'I', 'Low Risk')
Original Data
Gamma PDF k=6.90, theta=1113.96, n=190
Distibution for (1, '35-50', 'I', 'Low Risk')
Original Data
Gamma PDF k=6.48, theta=1321.41, n=250
Distibution for (0, '35-50', 'I', 'Moderate Risk')
Original Data
Gamma PDF k=7.19, theta=1000.04, n=344
Distibution for (1, '35-50', 'I', 'Moderate Risk')
Original Data
Gamma PDF k=8.38, theta=1000.06, n=741
Distibution for (0, '35-50', 'I', 'High Risk')
Original Data
Gamma PDF k=6.84, theta=1000.04, n=311
Distibution for (1, '35-50', 'I', 'High Risk')
Original Data
Gamma PDF k=8.23, theta=1000.06, n=424
Density
Baseline Stepcount

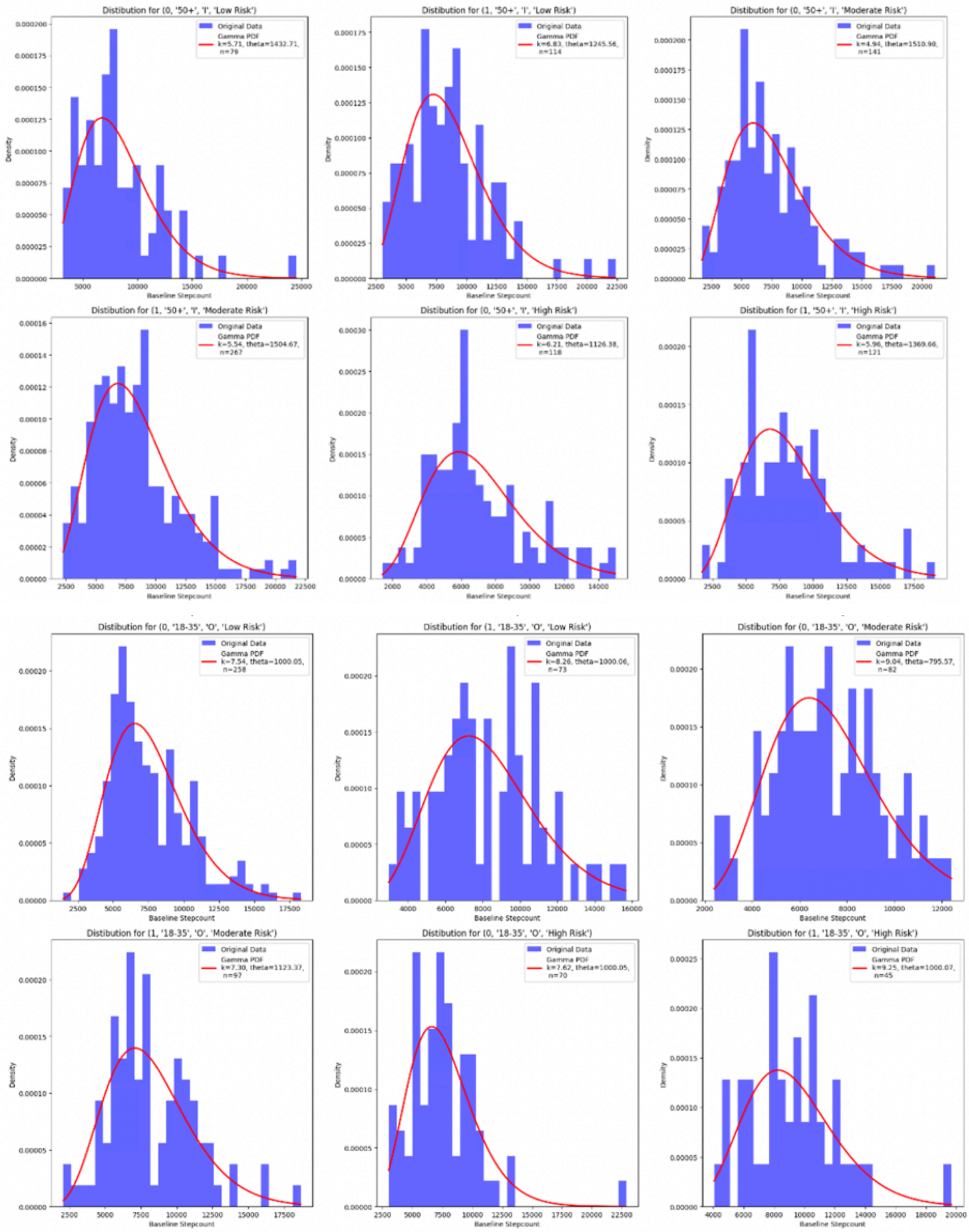
Distibution for (0, '50+', 'I', 'Low Risk')
Original Data
Gamma PDF
k=5.71, theta=1432.71, n=79
Distibution for (1, '50+', 'I', 'Low Risk')
Original Data
Gamma PDF
k=6.83, theta=1245.56, n=114
Distibution for (0, '50+', 'I', 'Moderate Risk')
Original Data
Gamma PDF
k=4.94, theta=1510.98, n=141
Distibution for (1, '50+', 'I', 'Moderate Risk')
Original Data
Gamma PDF
k=5.54, theta=1504.67, n=267
Distibution for (0, '50+', 'I', 'High Risk')
Original Data
Gamma PDF
k=6.21, theta=1126.38, n=118
Distibution for (1, '50+', 'I', 'High Risk')
Original Data
Gamma PDF
k=5.96, theta=1369.66, n=121
Distibution for (0, '18-35', 'O', 'Low Risk')
Original Data
Gamma PDF
k=7.54, theta=1000.05, n=258
Distibution for (1, '18-35', 'O', 'Low Risk')
Original Data
Gamma PDF
k=8.26, theta=1000.06, n=73
Distibution for (0, '18-35', 'O', 'Moderate Risk')
Original Data
Gamma PDF
k=9.04, theta=795.57, n=82
Distibution for (1, '18-35', 'O', 'Moderate Risk')
Original Data
Gamma PDF
k=7.30, theta=1123.37, n=97
Distibution for (0, '18-35', 'O', 'High Risk')
Original Data
Gamma PDF
k=7.62, theta=1000.05, n=70
Distibution for (1, '18-35', 'O', 'High Risk')
Original Data
Gamma PDF
k=9.25, theta=1000.07, n=45
Density
Baseline Stepcount

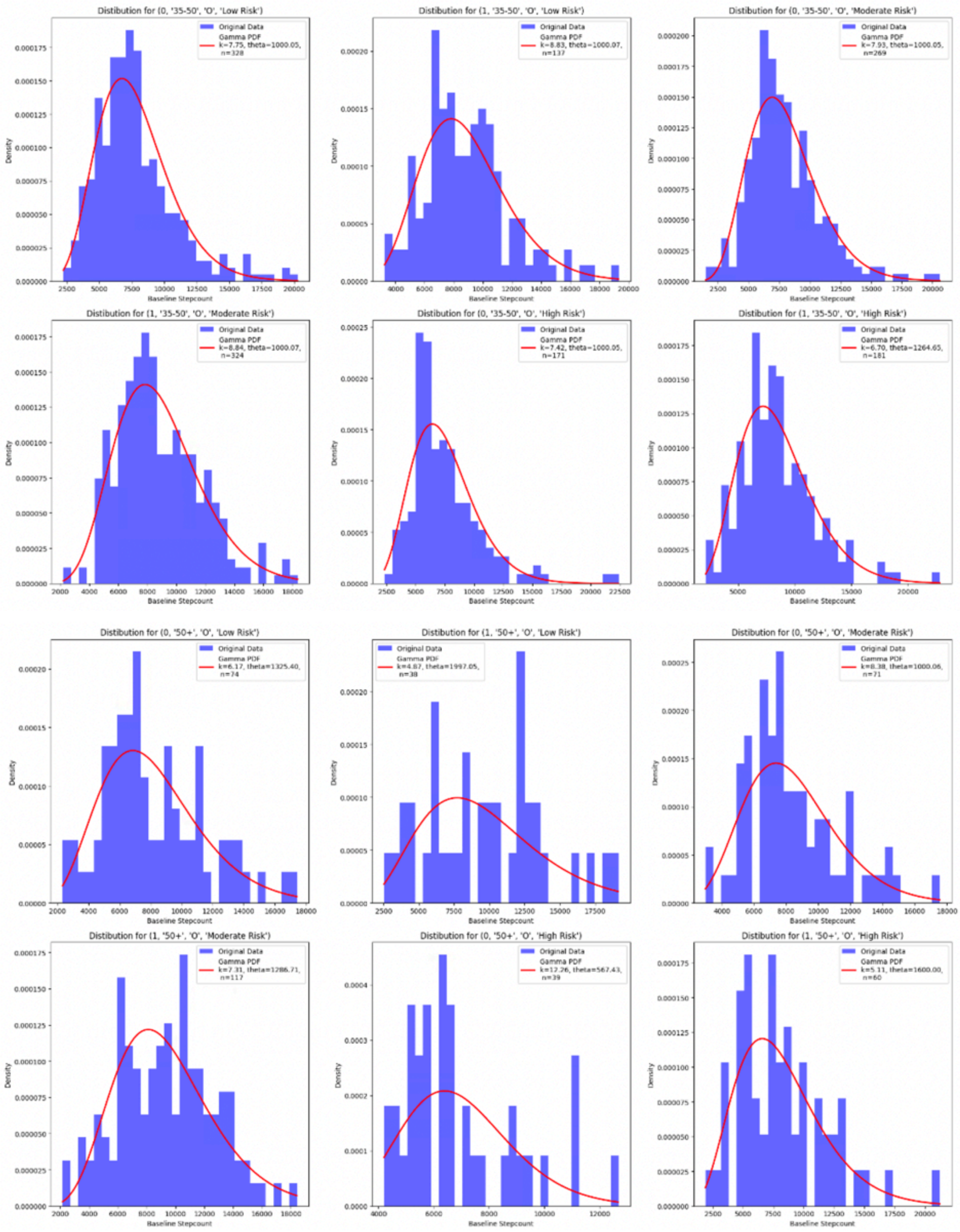


**Figure S8.** Distributions of baseline step count records for 72 strata of age group, ethnicity, sex and BMI group modelled with gamma functions using MLE.

**Table S2.** Table of shape and scale parameters of gamma distributions used to approximate baseline step count distributions by ethnicity, gender, age group and BMI risk group.

| Gamma distribution parameter by strata | | | Chinese | | Malay | | Indian | | Other | |
|---|---|---|---|---|---|---|---|---|---|---|
| | | | Male | Female | Male | Female | Male | Female | Male | Female |
| **Low Risk BMI group** | 18-30 | shape | 8.43 | 8.14 | 6.93 | 6.5 | 7.35 | 8.16 | 6.5 | 6.52 |
| | | scale | 1000.06 | 930.78 | 1291.63 | 1092.2 | 1136.4 | 895.66 | 1213.29 | 1166.32 |
| | 30-50 | shape | 7.04 | 6.59 | 6.79 | 6.45 | 6.82 | 6.15 | 8.83 | 7.02 |
| | | scale | 1203.07 | 1164.26 | 1248.43 | 1151.77 | 1322.4 | 1253.51 | 1000.07 | 1055.74 |
| | 50+ | shape | 5.69 | 5.64 | 4.18 | 4.36 | 6.28 | 6.12 | 5.99 | 6.6 |
| | | scale | 1623.19 | 1493.26 | 2082.16 | 1663.28 | 1353.44 | 1389.28 | 1825.67 | 1204.39 |
| **Moderate Risk BMI group** | 18-30 | shape | 8.6 | 7.63 | 8.83 | 7.57 | 7.83 | 5.95 | 10.02 | 8.02 |
| | | scale | 1000.06 | 1000.05 | 1000.07 | 917.64 | 1105.01 | 1266.35 | 855.89 | 888.13 |
| | 30-50 | shape | 8.36 | 7.22 | 8.18 | 6.9 | 8.44 | 7.34 | 8.72 | 9.23 |
| | | scale | 1000.06 | 1059.96 | 1000.06 | 1000.04 | 1000.06 | 1000.04 | 1000.07 | 846.05 |
| | 50+ | shape | 5.92 | 5.57 | 6.21 | 5.42 | 5.49 | 5.22 | 9.55 | 8.44 |
| | | scale | 1510.92 | 1477.99 | 1185.82 | 1354.5 | 1548.69 | 1474.78 | 1000.08 | 1000.06 |
| **High Risk BMI group** | 18-30 | shape | 8.4 | 7.54 | 7.63 | 8.14 | 7.27 | 6.55 | 12.32 | 9.21 |
| | | scale | 1000.06 | 1000.05 | 1107.56 | 841.59 | 1106.66 | 1061.73 | 697.61 | 771.11 |
| | 30-50 | shape | 7.51 | 6.99 | 6.79 | 6.66 | 8.32 | 6.73 | 6.62 | 6.37 |
| | | scale | 1114.76 | 1057.86 | 1126.98 | 1000.03 | 1000.06 | 1000.04 | 1255.11 | 1143.97 |
| | 50+ | shape | 6.07 | 5.43 | 5.95 | 5.2 | 6.59 | 5.52 | 5.82 | 10.63 |
| | | scale | 1411.87 | 1397.77 | 1441.67 | 1196.9 | 1218.01 | 1294.97 | 1405.12 | 656.52 |

We obtained participant's change in step count data by subtracting participant's baseline step counts from the daily average non-zero step count data averaged across the intervention period challenge weeks (weeks 12 - 37 in NSC3, NSC4 and NSC5). We used the data of only those actively enrolled participants that clocked a weekly average of at least 1000 daily steps averaged across 26 weeks of the challenge, yielding a total of 66,734 participants. Similar to baseline step counts, we modelled the change in step count across 72 strata as gamma fluctuations whose shape and scale parameters we obtained using the MLE as seen in Figure S9 and Table S3 below.

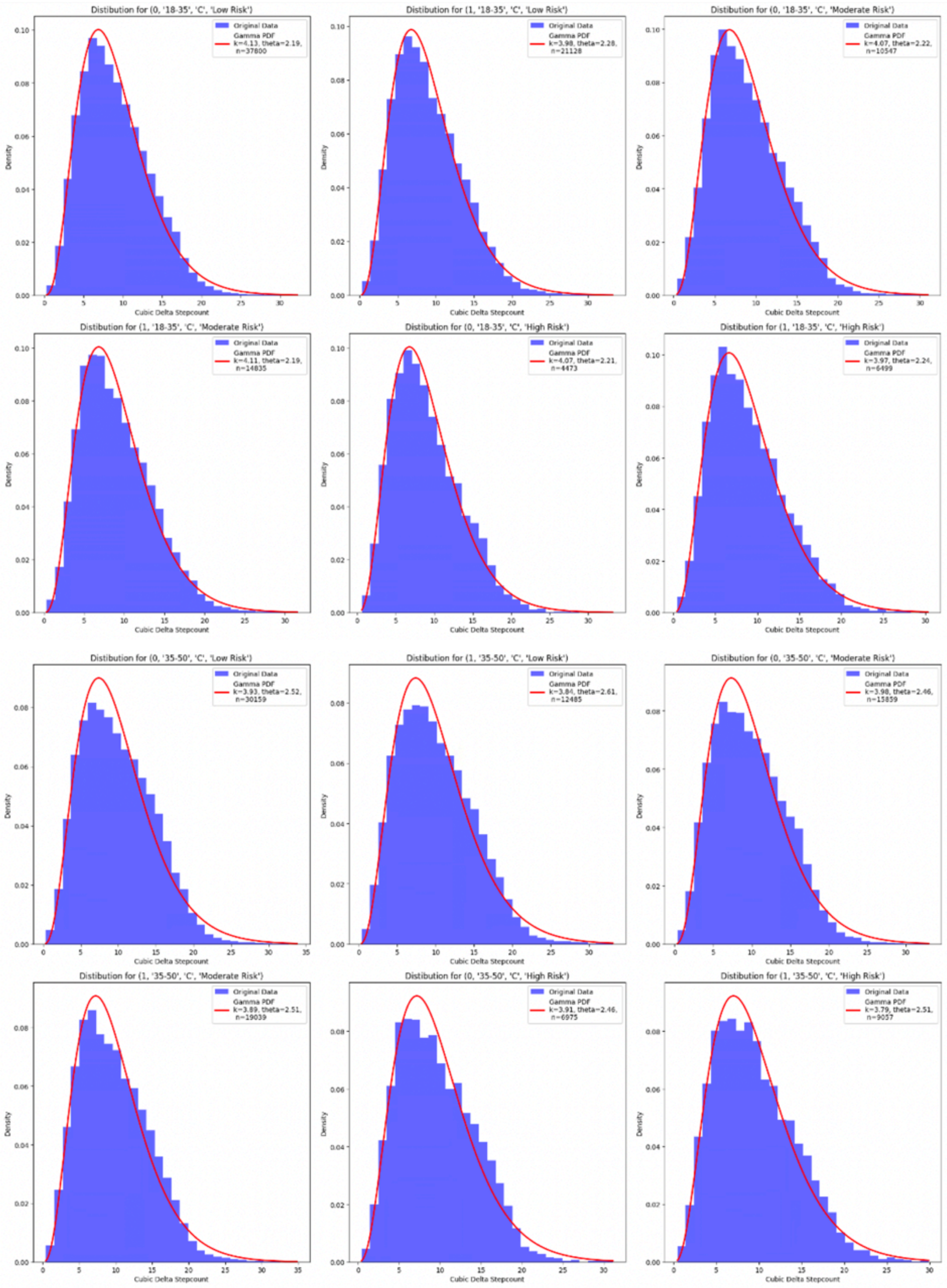
Distribution for (0, '18-35', 'C', 'Low Risk')
Original Data
Gamma PDF k=4.13, theta=2.19, n=37800
Distibution for (1, '18-35', 'C', 'Low Risk')
Original Data
Gamma PDF k=3.98, theta=2.28, n=21128
Distibution for (0, '18-35', 'C', 'Moderate Risk')
Original Data
Gamma PDF k=4.07, theta=2.22, n=10547
Distibution for (1, '18-35', 'C', 'Moderate Risk')
Original Data
Gamma PDF k=4.11, theta=2.19, n=14835
Distibution for (0, '18-35', 'C', 'High Risk')
Original Data
Gamma PDF k=4.07, theta=2.21, n=4473
Distibution for (1, '18-35', 'C', 'High Risk')
Original Data
Gamma PDF k=3.97, theta=2.24, n=6499
Distibution for (0, '35-50', 'C', 'Low Risk')
Original Data
Gamma PDF k=3.93, theta=2.52, n=30159
Distibution for (1, '35-50', 'C', 'Low Risk')
Original Data
Gamma PDF k=3.84, theta=2.61, n=12485
Distibution for (0, '35-50', 'C', 'Moderate Risk')
Original Data
Gamma PDF k=3.98, theta=2.46, n=15859
Distibution for (1, '35-50', 'C', 'Moderate Risk')
Original Data
Gamma PDF k=3.89, theta=2.51, n=19039
Distibution for (0, '35-50', 'C', 'High Risk')
Original Data
Gamma PDF k=3.91, theta=2.46, n=6975
Distibution for (1, '35-50', 'C', 'High Risk')
Original Data
Gamma PDF k=3.79, theta=2.51, n=9057
Density
Cubic Delta Stepcount

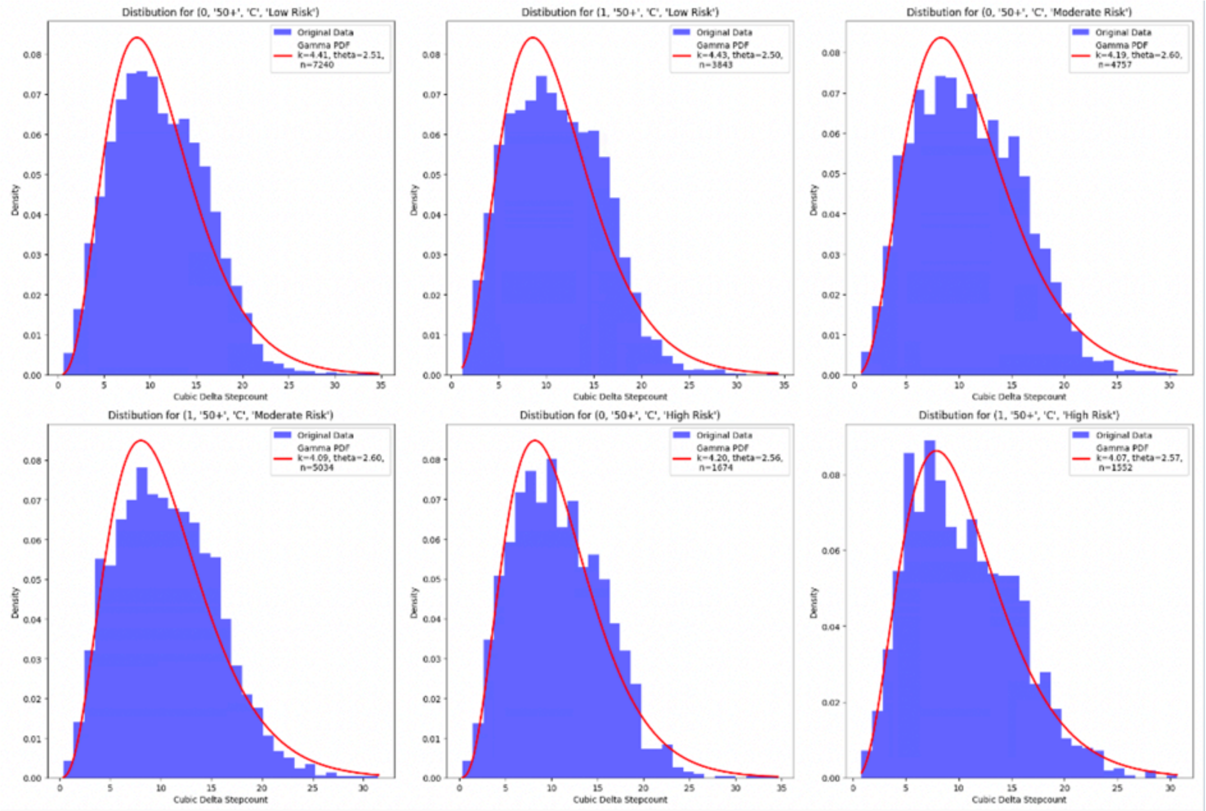
Distibution for (0, '50+', 'C', 'Low Risk')
Original Data
Gamma PDF
k=4.41, theta=2.51,
n=7240
Cubic Delta Stepcount
Density
Distibution for (1, '50+', 'C', 'Low Risk')
Original Data
Gamma PDF
k=4.43, theta=2.50,
n=3843
Cubic Delta Stepcount
Density
Distibution for (0, '50+', 'C', 'Moderate Risk')
Original Data
Gamma PDF
k=4.19, theta=2.60,
n=4757
Cubic Delta Stepcount
Density
Distibution for (1, '50+', 'C', 'Moderate Risk')
Original Data
Gamma PDF
k=4.09, theta=2.60,
n=5034
Cubic Delta Stepcount
Density
Distibution for (0, '50+', 'C', 'High Risk')
Original Data
Gamma PDF
k=4.20, theta=2.56,
n=1674
Cubic Delta Stepcount
Density
Distibution for (1, '50+', 'C', 'High Risk')
Original Data
Gamma PDF
k=4.07, theta=2.57,
n=1552
Cubic Delta Stepcount
Density

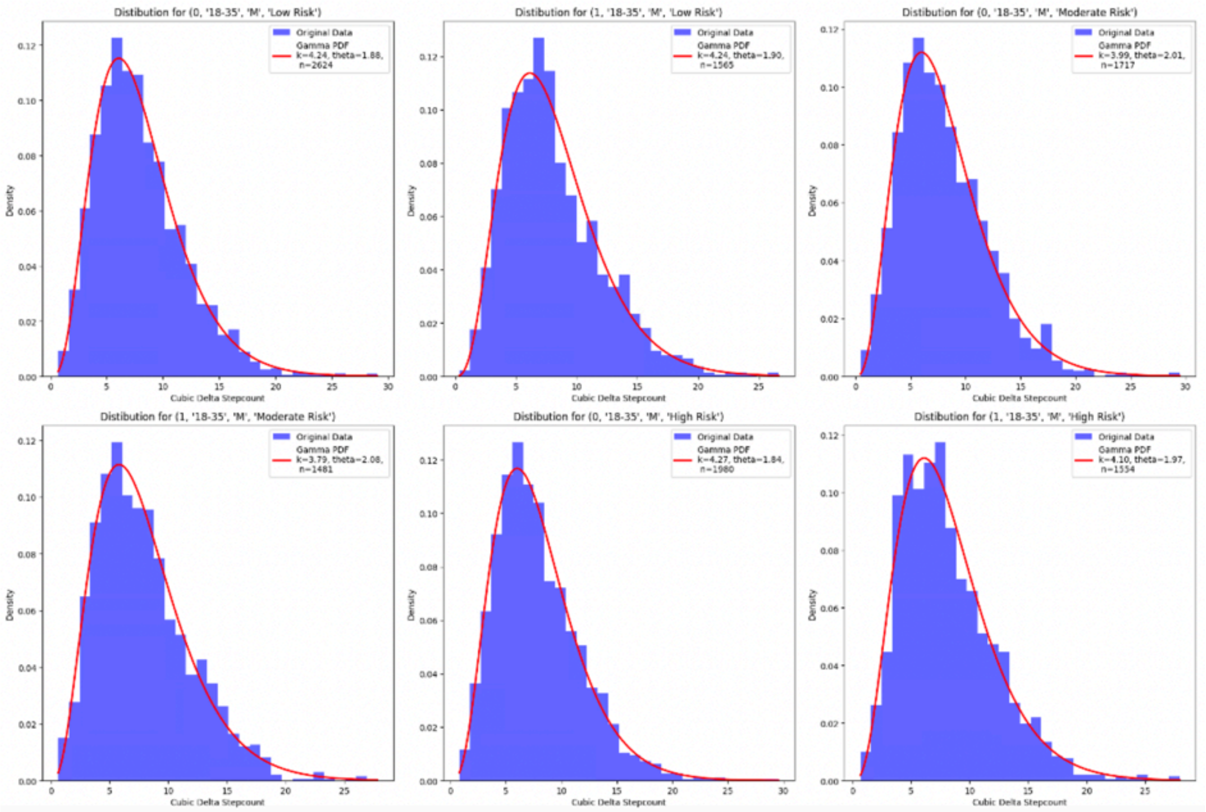
Distibution for (0, '18-35', 'M', 'Low Risk')
Original Data
Gamma PDF
k=4.24, theta=1.88,
n=2624
Cubic Delta Stepcount
Density
Distibution for (1, '18-35', 'M', 'Low Risk')
Original Data
Gamma PDF
k=4.24, theta=1.90,
n=1565
Cubic Delta Stepcount
Density
Distibution for (0, '18-35', 'M', 'Moderate Risk')
Original Data
Gamma PDF
k=3.99, theta=2.01,
n=1717
Cubic Delta Stepcount
Density
Distibution for (1, '18-35', 'M', 'Moderate Risk')
Original Data
Gamma PDF
k=3.79, theta=2.08,
n=1481
Cubic Delta Stepcount
Density
Distibution for (0, '18-35', 'M', 'High Risk')
Original Data
Gamma PDF
k=4.27, theta=1.84,
n=1980
Cubic Delta Stepcount
Density
Distibution for (1, '18-35', 'M', 'High Risk')
Original Data
Gamma PDF
k=4.10, theta=1.97,
n=1554
Cubic Delta Stepcount
Density

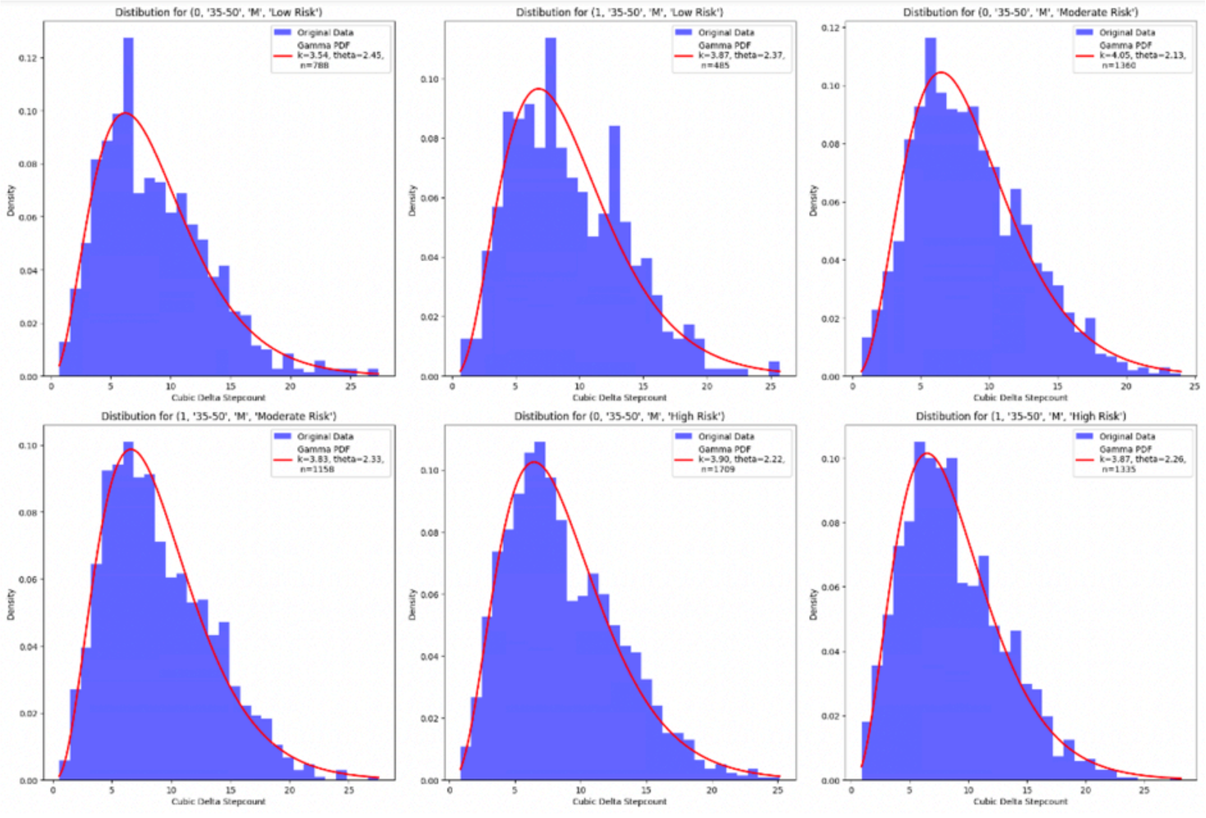
Distibution for (0, '35-50', 'M', 'Low Risk')
Original Data
Gamma PDF
k=3.54, theta=2.45,
n=788
Density
Cubic Delta Stepcount
Distibution for (1, '35-50', 'M', 'Low Risk')
Original Data
Gamma PDF
k=3.87, theta=2.37,
n=485
Density
Cubic Delta Stepcount
Distibution for (0, '35-50', 'M', 'Moderate Risk')
Original Data
Gamma PDF
k=4.05, theta=2.13,
n=1360
Density
Cubic Delta Stepcount
Distibution for (1, '35-50', 'M', 'Moderate Risk')
Original Data
Gamma PDF
k=3.83, theta=2.33,
n=1158
Density
Cubic Delta Stepcount
Distibution for (0, '35-50', 'M', 'High Risk')
Original Data
Gamma PDF
k=3.90, theta=2.22,
n=1709
Density
Cubic Delta Stepcount
Distibution for (1, '35-50', 'M', 'High Risk')
Original Data
Gamma PDF
k=3.87, theta=2.26,
n=1335
Density
Cubic Delta Stepcount

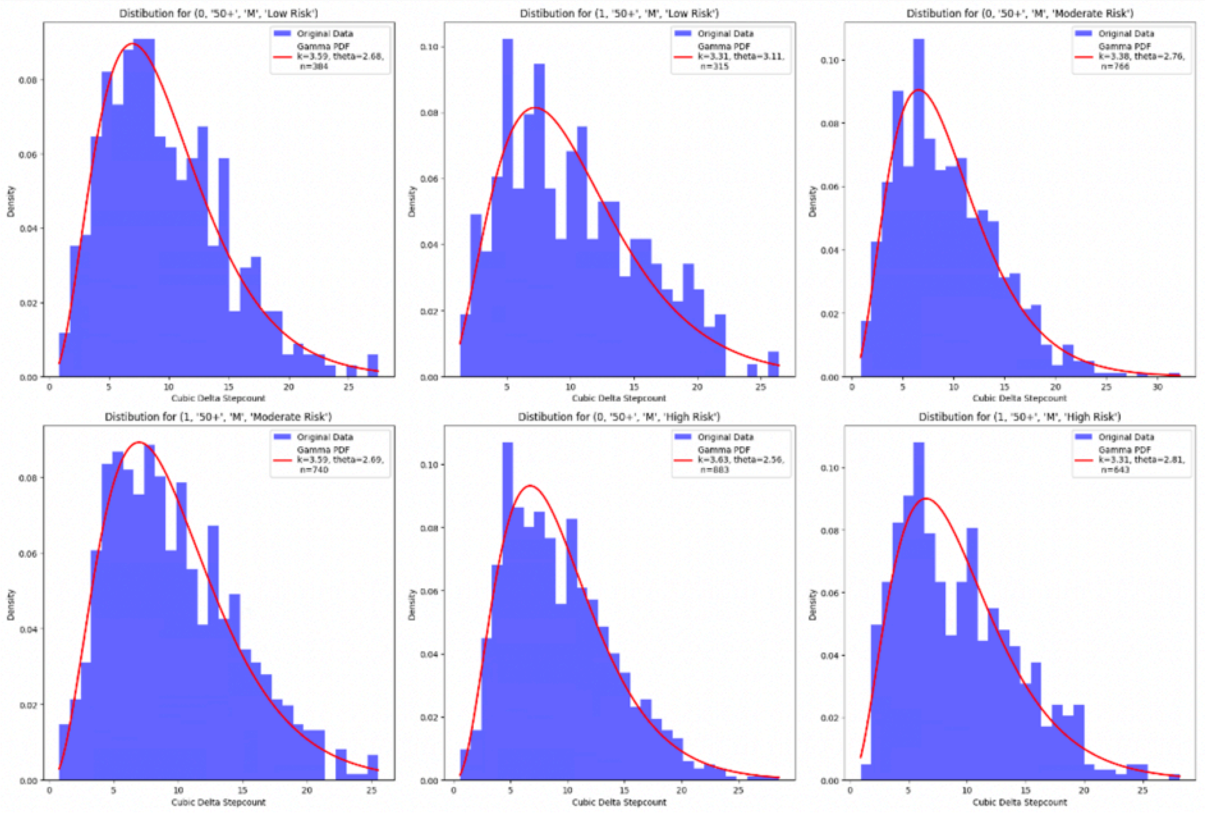
Distibution for (0, '50+', 'M', 'Low Risk')
Original Data
Gamma PDF
k=3.59, theta=2.68,
n=384
Density
Cubic Delta Stepcount
Distibution for (1, '50+', 'M', 'Low Risk')
Original Data
Gamma PDF
k=3.31, theta=3.11,
n=315
Density
Cubic Delta Stepcount
Distibution for (0, '50+', 'M', 'Moderate Risk')
Original Data
Gamma PDF
k=3.38, theta=2.76,
n=766
Density
Cubic Delta Stepcount
Distibution for (1, '50+', 'M', 'Moderate Risk')
Original Data
Gamma PDF
k=3.59, theta=2.69,
n=740
Density
Cubic Delta Stepcount
Distibution for (0, '50+', 'M', 'High Risk')
Original Data
Gamma PDF
k=3.63, theta=2.56,
n=883
Density
Cubic Delta Stepcount
Distibution for (1, '50+', 'M', 'High Risk')
Original Data
Gamma PDF
k=3.31, theta=2.81,
n=643
Density
Cubic Delta Stepcount

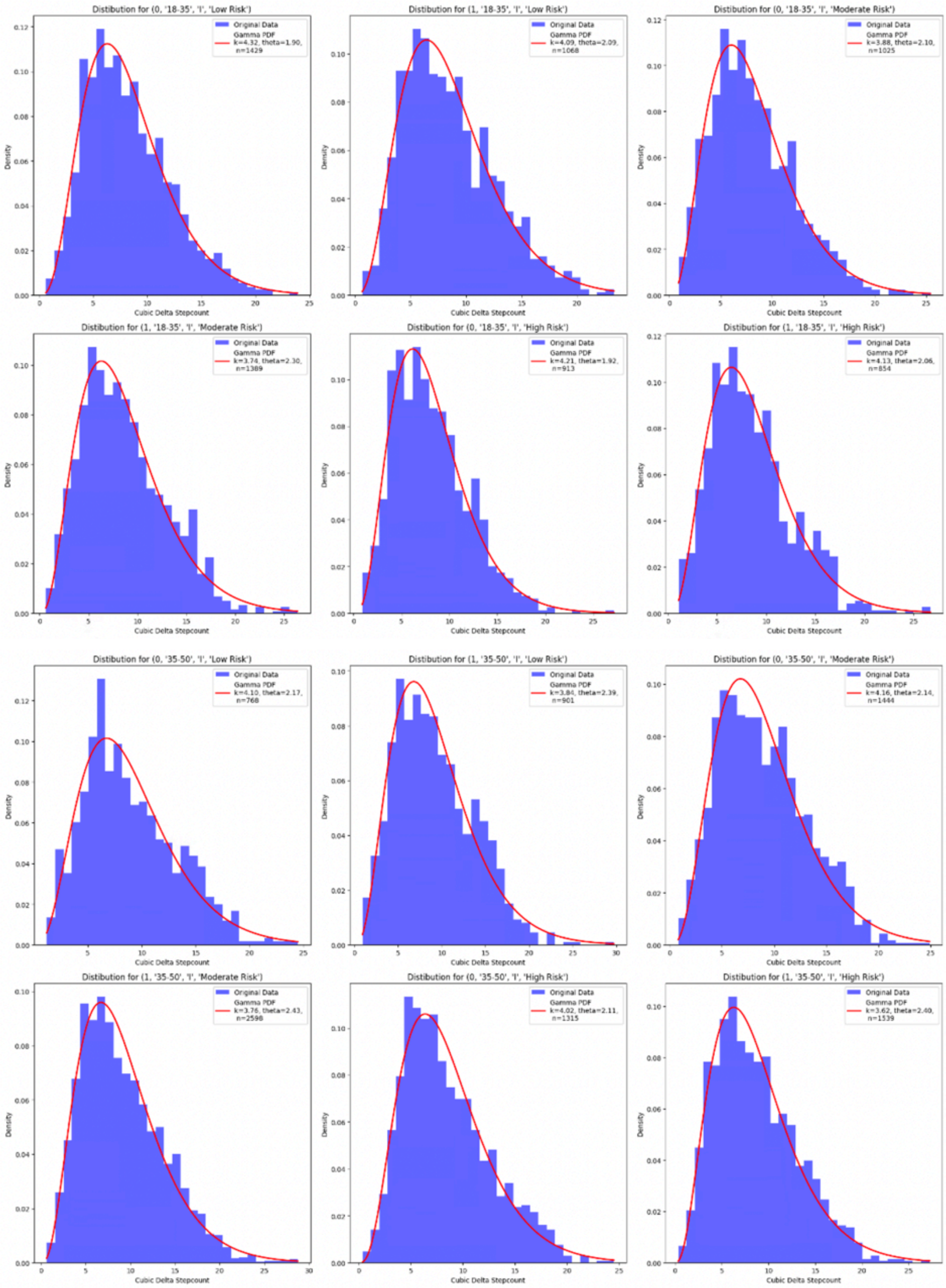
Distibution for (0, '18-35', 'I', 'Low Risk')
Original Data
Gamma PDF
k=4.32, theta=1.90, n=1429
Distibution for (1, '18-35', 'I', 'Low Risk')
Original Data
Gamma PDF
k=4.09, theta=2.09, n=1068
Distibution for (0, '18-35', 'I', 'Moderate Risk')
Original Data
Gamma PDF
k=3.88, theta=2.10, n=1025
Distibution for (1, '18-35', 'I', 'Moderate Risk')
Original Data
Gamma PDF
k=3.74, theta=2.30, n=1389
Distibution for (0, '18-35', 'I', 'High Risk')
Original Data
Gamma PDF
k=4.21, theta=1.92, n=913
Distibution for (1, '18-35', 'I', 'High Risk')
Original Data
Gamma PDF
k=4.13, theta=2.06, n=854
Distribution for (0, '35-50', 'I', 'Low Risk')
Original Data
Gamma PDF
k=4.10, theta=2.17, n=768
Distibution for (1, '35-50', 'I', 'Low Risk')
Original Data
Gamma PDF
k=3.84, theta=2.39, n=901
Distibution for (0, '35-50', 'I', 'Moderate Risk')
Original Data
Gamma PDF
k=4.16, theta=2.14, n=1444
Distibution for (1, '35-50', 'I', 'Moderate Risk')
Original Data
Gamma PDF
k=3.76, theta=2.43, n=2598
Distibution for (0, '35-50', 'I', 'High Risk')
Original Data
Gamma PDF
k=4.02, theta=2.11, n=1315
Distibution for (1, '35-50', 'I', 'High Risk')
Original Data
Gamma PDF
k=3.62, theta=2.40, n=1539
Density
Cubic Delta Stepcount

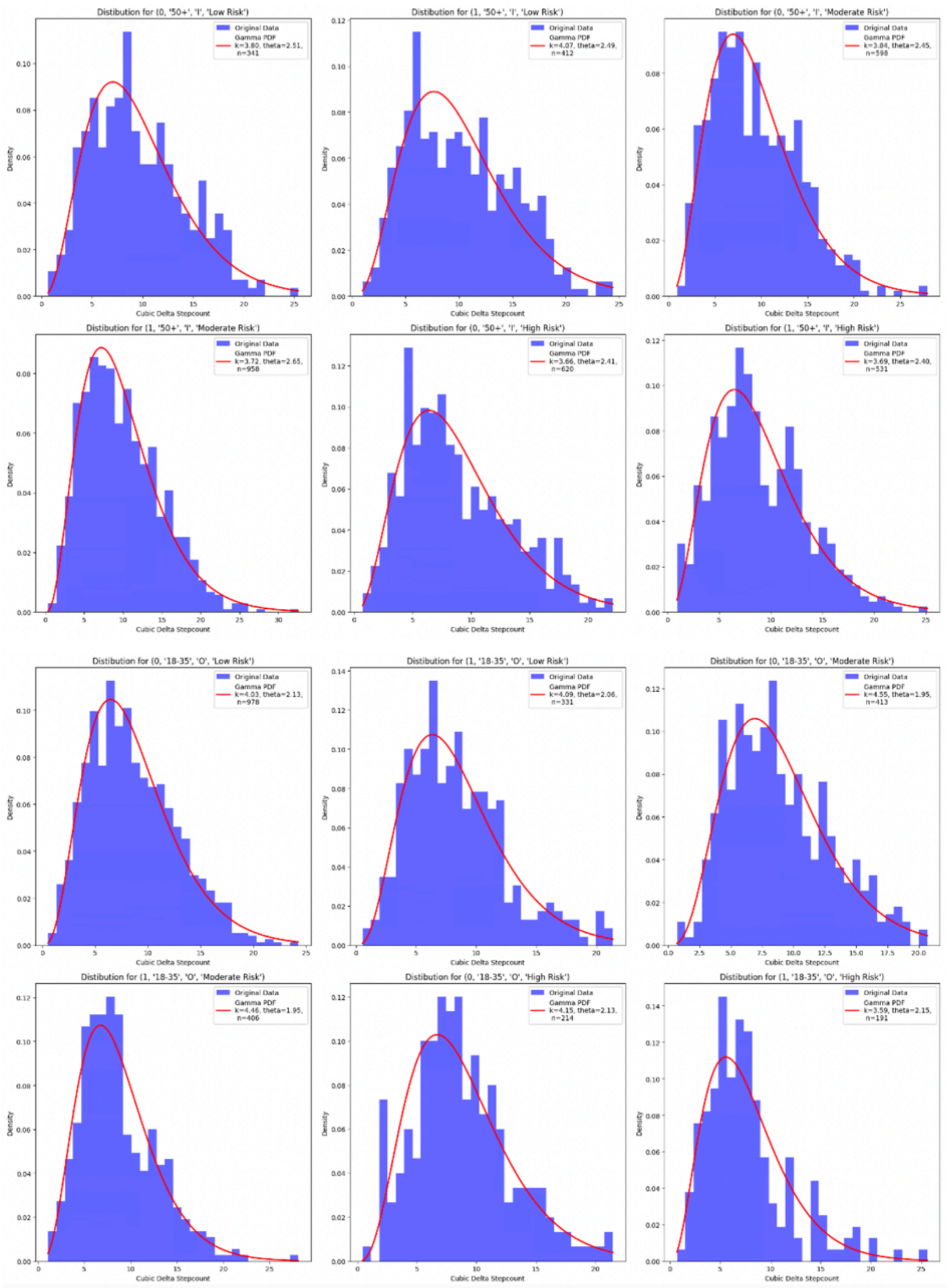
Distibution for (0, '50+', 'I', 'Low Risk')
Original Data
Gamma PDF k=3.80, theta=2.51, n=341
Distibution for (1, '50+', 'I', 'Low Risk')
Original Data
Gamma PDF k=4.07, theta=2.49, n=412
Distibution for (0, '50+', 'I', 'Moderate Risk')
Original Data
Gamma PDF k=3.84, theta=2.45, n=598
Distibution for (1, '50+', 'I', 'Moderate Risk')
Original Data
Gamma PDF k=3.72, theta=2.65, n=958
Distibution for (0, '50+', 'I', 'High Risk')
Original Data
Gamma PDF k=3.66, theta=2.41, n=620
Distibution for (1, '50+', 'I', 'High Risk')
Original Data
Gamma PDF k=3.69, theta=2.40, n=531
Distibution for (0, '18-35', 'O', 'Low Risk')
Original Data
Gamma PDF k=4.03, theta=2.13, n=978
Distibution for (1, '18-35', 'O', 'Low Risk')
Original Data
Gamma PDF k=4.09, theta=2.06, n=331
Distibution for (0, '18-35', 'O', 'Moderate Risk')
Original Data
Gamma PDF k=4.55, theta=1.95, n=413
Distibution for (1, '18-35', 'O', 'Moderate Risk')
Original Data
Gamma PDF k=4.46, theta=1.95, n=406
Distibution for (0, '18-35', 'O', 'High Risk')
Original Data
Gamma PDF k=4.15, theta=2.13, n=214
Distibution for (1, '18-35', 'O', 'High Risk')
Original Data
Gamma PDF k=3.59, theta=2.15, n=191
Density
Cubic Delta Stepcount

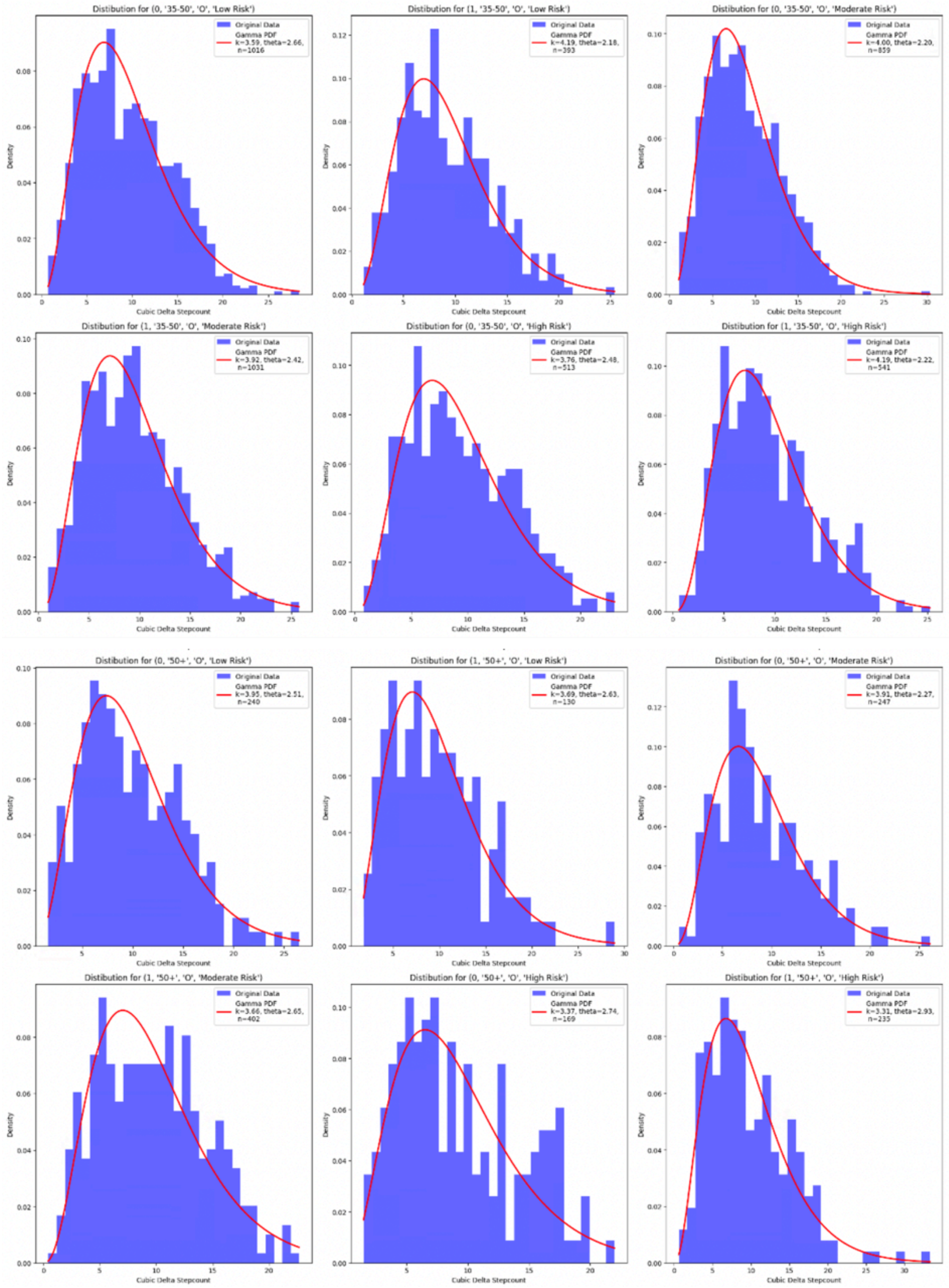


**Figure S9.** Distributions of change in step count records for 72 strata of age group, ethnicity, gender and BMI group modelled with gamma functions using MLE.

**Table S3.** Table of shape and scale parameters of gamma distributions used to approximate cubic root of the change in step count distributions ethnicity, gender, age group and BMI risk group.

| Gamma distribution parameter by strata | | | Chinese | | Malay | | Indian | | Other | |
|---|---|---|---|---|---|---|---|---|---|---|
| | | | Male | Female | Male | Female | Male | Female | Male | Female |
| **Low Risk BMI group** | 18-30 | shape | 3.93 | 4.08 | 4.13 | 4.17 | 4.14 | 4.34 | 4.26 | 4.08 |
| | | scale | 2.35 | 2.26 | 1.96 | 1.97 | 2.06 | 1.91 | 2.07 | 2.16 |
| | 30-50 | shape | 3.82 | 3.86 | 3.57 | 3.63 | 3.52 | 3.67 | 3.49 | 3.93 |
| | | scale | 2.66 | 2.62 | 2.52 | 2.44 | 2.46 | 2.38 | 2.66 | 2.53 |
| | 50+ | shape | 3.69 | 3.74 | 3.44 | 3.72 | 3.75 | 3.75 | 3.52 | 4.19 |
| | | scale | 2.99 | 2.92 | 3.13 | 2.6 | 2.62 | 2.45 | 2.64 | 2.32 |
| **Moderate Risk BMI group** | 18-30 | shape | 3.97 | 4.08 | 3.75 | 4.5 | 4.02 | 3.98 | 4.09 | 3.95 |
| | | scale | 2.29 | 2.26 | 2.18 | 1.81 | 2.14 | 2.05 | 2.03 | 2.28 |
| | 30-50 | shape | 3.8 | 3.96 | 3.78 | 4.05 | 3.48 | 4.14 | 4.01 | 4 |
| | | scale | 2.6 | 2.52 | 2.31 | 2.19 | 2.6 | 2.14 | 2.37 | 2.24 |
| | 50+ | shape | 3.61 | 3.72 | 3.86 | 3.44 | 3.59 | 3.99 | 3.5 | 3.46 |
| | | scale | 2.96 | 2.88 | 2.54 | 2.68 | 2.67 | 2.32 | 2.55 | 2.54 |
| **High Risk BMI group** | 18-30 | shape | 3.89 | 3.97 | 4.3 | 4.2 | 3.87 | 3.85 | 3.95 | 4.39 |
| | | scale | 2.32 | 2.28 | 1.81 | 1.88 | 2.2 | 2.05 | 1.99 | 2.06 |
| | 30-50 | shape | 3.72 | 3.78 | 3.88 | 3.74 | 3.61 | 3.73 | 4.19 | 3.71 |
| | | scale | 2.58 | 2.59 | 2.23 | 2.27 | 2.43 | 2.25 | 2.27 | 2.65 |
| | 50+ | shape | 3.66 | 3.81 | 3.26 | 3.81 | 3.67 | 3.6 | 4.18 | 3.03 |
| | | scale | 2.85 | 2.8 | 2.83 | 2.47 | 2.37 | 2.46 | 2.39 | 2.91 |

### Supplementary Information 2. NSC participation models.

In this section we explain how NSC participation is simulated in DEMOS. Every year starting from 2015, the year of the first NSC season, a random number between 0 and 1 is drawn from a uniform distribution for each adult in our microsimulation. If this random number is less than the associated probability of NSC participation, the individual is considered to have been an NSC participant in that year and the year in which this occurs is recorded as the year of NSC participation for the individual. If the threshold is not met, the individual is not recorded as an NSC participant in the given year. Probability of NSC participation in the given year depends on whether the individual was an NSC participant in the previous. For those that were, the probability of NSC participation in the current year is defined by the retention rate, while for those that were not, it is defined by the enrolment rate. These rates are based on the past NSC participation data.

Enrolment rate is the percentage of the SG population that enrolled actively into the NSC programme in the current year and did not participate in the previous year. Retention rate is the percentage of the current year's active NSC participants that remained active in the subsequent season. We also define dropout rate, which is a complement of the retention rate, i.e., retention rate + dropout rate = 1. In other words, it is the percentage of the current year's active NSC participants that did not participate in NSC in the subsequent year. Tables S4 and S5 show the rates used in our microsimulation. Table S4 shows past rates based on the NSC participation data, while Table S5 shows the rates used in 5 different NSC scenarios, which are differentiated only by the variability in the coverage and effectiveness of NSC enrolment and retention.

**Table S4.** Past enrolment, retention and dropout rates per year/NSC season.

| Year/NSC Season | Enrolment rate | Retention rate | Dropout rate |
|---|---|---|---|
| 2015/NSC1 | 3.0% | 12.6% | 87.4% |
| 2016/NSC2 | 6.9% | 5.8% | 94.2% |
| 2017/NSC3 | 11.7% | 22.9% | 77.1% |
| 2018/NSC4 | 9.1% | 32.4% | 67.6% |
| 2019/NSC5 | 7.9% | 29.6% | 70.4% |

**Table S5.** Enrolment rate, retention rate and rationale for five simulation scenarios covering years 2022-2050.

| Simulation parameter | No NSC | Average NSC | Peak NSC | Double NSC | Full NSC |
|---|---|---|---|---|---|
| Enrolment rate | 0% | 9.6% | 11.7% | 19.2% | 100% |
| Retention rate | 0% | 28.3% | 32.4% | 56.6% | 100% |

The baseline scenario ("No NSC") assumes the termination of the NSC programme from 2022 onwards and allows us to quantify the underlying burden of obesity and T2DM in the absence of any further population-wide physical activity intervention. Including this scenario is essential for establishing a reference point against which the incremental benefits of any public health programme can be rigorously assessed. The "Average NSC" scenario projects future enrolment and retention at the mean rates observed in NSC3–5 (enrolment: 9.6%, retention: 28.3%), representing a realistic continuation of past practice. The "Peak NSC" scenario models the highest rates achieved to date (enrolment: 11.7%, retention: 32.4%), capturing the programme's most successful implementation. Importantly, we included a "Double NSC" scenario, which assumes a doubling of both enrolment and retention relative to the Average NSC (enrolment: 19.2%, retention: 56.6%). This scenario is designed to test the public health impact of ambitious, though potentially attainable, expansion under intensified policy commitment, increased resource allocation, and enhanced community engagement strategies. It provides key insights into the upper range of feasible outcomes that could be targeted by scaling up intervention efforts. Finally, the "Full NSC" scenario assumes universal enrolment and retention among all eligible adults, representing a theoretical maximum for intervention coverage. While this is not a realistic expectation for voluntary health interventions, modelling the Full NSC scenario is valuable for understanding the ceiling of potential benefit and for quantifying the full scope of preventable disease burden attributable to insufficient physical activity. Collectively, these scenarios enable a nuanced, policy-relevant analysis of both the incremental and maximal potential of large-scale physical activity programmes to address the growing burden of obesity and diabetes at the population level.

Figure S10 below shows NSC participation model flow design. In any given year, the NSC participants can transition out of the "First Time", "Continuing" and "Drop out" pools. New NSC participants, designated as those that did not participate in NSC in the previous year, enter the "First Time" pool if they meet the enrolment threshold. The same was performed for those that are already active NSC participants, where they may continue or drop out, based on the associated retention rates. So every year starting from 2016 we have individuals that joined NSC for the first time, those that stayed in the NSC and those that quit or never enrolled.

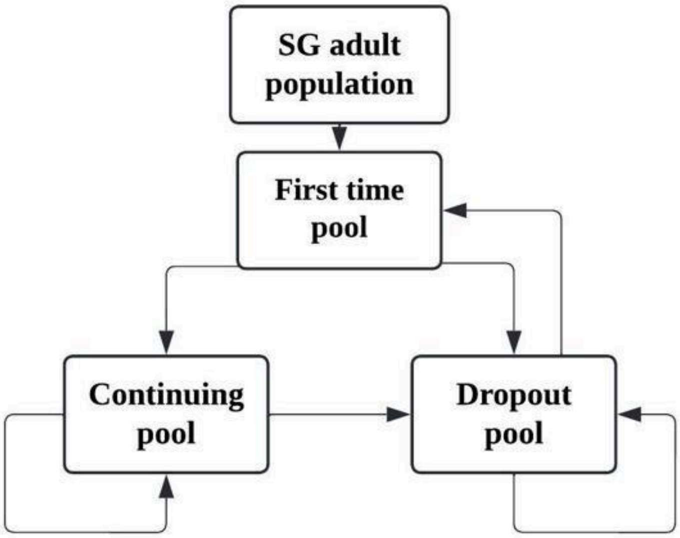


**Figure S10.** NSC participation model flow.

**First time participant model**

To estimate the effect of the NSC programme on body mass index (BMI), we modelled the change in BMI ($\Delta BMI$ in $kg/m^2$) during the first year of participation using a Bayesian generalized linear model (GLM) with an identity link. Predictor variables included sex (binary indicator for male, $g$), age in years ($a$), baseline BMI ($c$), the cubic root of baseline step count ($b$), the cubic root of change in step count ($d$), and indicator variables for Indian ($I$) and Malay ($M$) ethnicity. The cubic root transformation was applied to step count variables to address right-skewness and improve linearity. Standard normal priors ($\beta_j \sim N(0, 1)\ for\ j \in [0, 7]$) were specified for all regression coefficients, and a half-Cauchy prior for the residual standard deviation ($\sigma \sim Half - Cauchy(0, 2.5)$). Model specification was as follows:

$\Delta BMI_i \sim N(\mu_i, \sigma)$

$\mu_i = \beta_0 + \beta_1 g_i + \beta_2 a_i + \beta_3 c_i + \beta_4 b_i + \beta_5 d_i + \beta_6 I_i + \beta_7 M_i$

where $g_i$ indicates where the individual is male (binary: 0/1), $a_i$ denotes age (in years), $c_i$ indicates current BMI (kg/m²), $b_i$ represents the cubic root of baseline step count, $d_i$ denotes the cubic root of change in step count, $I_i$ indicates Indian ethnicity (binary: 0/1) and $M_i$ represents Malay ethnicity (binary: 0/1).

Posterior distributions for all model parameters were obtained by MCMC sampling using 5000 MCMC draws with the first 10% of MCMC iterations discarded as burn-in. We assessed convergence by inspection of each trace plot.

For use in the subsequent microsimulation, point estimates (posterior means) of the regression coefficients were calculated, resulting in the following model specification for the predicted change in BMI for individual $i$:

$$\Delta BMI_i = 3.3152 + 0.1495g_i - 0.0039a_i - 0.1263c_i - 0.0124b_i - 0.0106d_i + 0.1581I_i + 0.4592M_i$$

Model development and validation were conducted using data from 66,734 first-time NSC participants from NSC3–5 with no prior programme enrolment. To ensure data quality and model validity, the datasets were split into training, testing and validation sets and comprehensive diagnostic testing was performed. Residuals were examined for normality (Q-Q plots) and homoscedasticity; multicollinearity was assessed using variance inflation factors (VIF) for all predictors. Model selection was based on the lowest Bayesian Information Criterion (BIC) across alternative specifications, including models with different variable transformations and interaction terms. Posterior predictive checks were performed to evaluate model fit, and predictive accuracy was summarised using the posterior distributions of mean absolute error (MAE) and mean absolute percentage error (MAPE). Statistical support for each coefficient was assessed by verifying that the corresponding 95% Bayesian credible intervals excluded zero. Overall, the model demonstrated robust performance and stability across multiple posterior predictive validation sets.

**Continuing participant model**

We modelled change in the NSC participants' BMI ($\Delta BMI$) in the consecutive year of participation using Bayesian GLM with identity link. Predictor variables included gender (binary indicator for male, $g$), age in years ($a$), baseline BMI ($c$), the cubic root of baseline step count ($b$), the cubic root of change in step count ($d$), indicator variables for Indian ($I$) and Malay ($M$) ethnicity and change in the participant's BMI in the previous year($\Delta BMI^p$). The cubic root transformation was applied to step count variables to address right-skewness and improve linearity. Standard normal priors ($\beta_j \sim N(0, 1)\ for\ j \in [0, 8]$) were specified for all regression coefficients, and a half-Cauchy prior for the residual standard deviation ($\sigma \sim Half - Cauchy(0, 2.5)$). Model specification was as follows:

$$\Delta BMI_i \sim N(\mu_i, \sigma),$$

$$\mu_i = \beta_0 + \beta_1 g_i + \beta_2 a_i + \beta_3 c_i + \beta_4 b_i + \beta_5 d_i + \beta_6 I_i + \beta_7 M_i + \beta_8 \Delta BMI^p_i$$

where $\Delta BMI^p_i$ denotes change in BMI in the previous year (kg/m²), $g_i$ represents male gender (binary: 0/1), $a_i$ denotes age (in years), $c_i$ indicates current BMI (kg/m²), $d_i$ denotes the cubic root of change in step count, $I_i$ indicates Indian ethnicity (binary: 0/1) and $M_i$ represents Malay ethnicity (binary: 0/1).

Posterior distributions for all model parameters were obtained by Markov chain Monte Carlo (MCMC) sampling using 5000 MCMC draws with the first 10% of MCMC iterations discarded as burn-in. We assessed convergence by inspection of each trace plot.

For use in the subsequent microsimulation, point estimates (posterior means) of the regression coefficients were extracted, resulting in the following model specification for the predicted change in BMI for individual $i$:

$$\Delta BMI_i = 3.1269 + 0.1191g_i - 0.0101a_i - 0.1121c_i - 0.0101d_i + 0.1251I_i + 0.2871M_i - 0.0695\Delta BMI^p_i$$

Model development and validation were conducted using data from 25,966 continuing NSC participants from NSC3-5 who were also a part of the NSC participant pool in their respective previous season. To ensure data quality and model validity, the datasets were split into training, testing and validation sets and comprehensive diagnostic testing was performed. Residuals were examined for normality (Q-Q plots) and homoscedasticity; multicollinearity was assessed using variance inflation factors (VIF) for all predictors. Model selection was based on the lowest Bayesian Information Criterion (BIC) across alternative specifications, including models with different variable transformations and interaction terms. Posterior predictive checks were performed to evaluate model fit, and predictive accuracy was summarised using the posterior distributions of mean absolute error (MAE) and mean absolute percentage error (MAPE). Statistical support for each coefficient was assessed by verifying that the corresponding 95% Bayesian credible intervals excluded zero. Overall, the model demonstrated robust performance and stability across multiple posterior predictive validation sets.
